# From Output Errors to Workflow Harm: A Practitioner-Audit Method for LLM-Mediated Research

**DOI:** 10.64898/2026.08.13.26360414

**Authors:** Davis Austria, Byrron McCollister, Jeremy E. Lindsey, Micheal Arowolo, Marian Okon

**Affiliations:** MS Health Informatics Program, Xavier University of Louisiana, New Orleans, LA, USA

**Keywords:** Large language models, workflow harm, practitioner audit, claimed verification, medical informatics

## Abstract

**Objective:** Formal large language model (LLM) evaluations score isolated prompts, but clinicians and health-informatics researchers meet model failures inside multi-step workflows where erroneous output can alter procedures or contaminate documents. We present TRACE (Tracking Reliability of AI-generated Conversational Evidence), a practitioner-audit framework for evaluating the downstream workflow reliability of conversational AI.

**Materials and Methods:** A method paper with an empirical demonstration: 45 documentation-positive incidents recorded by one clinician-informatician across scholarly, clinical informatics, and clinical-adjacent workflows over seven weeks, coded with a consequence-based severity rubric, an error definition, a taxonomy crosswalk, and a Response-Audit Scorecard. Three reviewer-authors independently coded a 16-incident subsample; three vendor-blinded AI comparators applied the taxonomy to all 45 incidents.

**Results:** Four categories tied as most frequent: verification failure, factual numerical error, tool-behavior misunderstanding, and citation or reference formatting (n=7 each). Four workflow-harm patterns recurred— procedural propagation, documentary contamination, trust-calibration disruption, and user-borne corrective burden—and one incident carried an estimated $2500 impact. Category agreement across three human reviewer-authors was low (Fleiss κ=0.155), whereas three AI comparators agreed substantially (Fleiss κ=0.632), suggesting taxonomy legibility under standardized conditions even where human judgment diverged.

**Discussion:** Category assignment is comparatively legible, whereas severity and claimed-verification remain judgment-dependent. The claimed-verification gap is a measurable failure mode distinct from hallucination, sycophancy, and over-refusal.

**Conclusion:** Practitioner audits with structured response scoring complement benchmarks by documenting workflow harm as an applied evaluation unit for clinical informatics and public-health work; this is a pilot that motivates, not estimates, error rates or cross-model comparisons.

## 1. Introduction

Large language model (LLM) evaluation has matured rapidly. Surveys distinguish factuality from faithfulness hallucinations,[1] and system cards add deployment categories such as harmful content, privacy, overreliance, and refusal.[2,3] Long-context studies show evidence may be underused even when present.[4] Calibration and U-Sophistry research examine when wrongness becomes more convincing under preference optimization,[5,6] and sycophancy work documents answer-shifting under user pressure.[7]

These taxonomies share a unit of analysis: the prompt or benchmark item. In practice, the same wrong answer can change a screening procedure, alter a manuscript, or require external review to prevent downstream damage. The US National Institute of Standards and Technology (NIST) AI Risk Management Framework emphasizes that artificial intelligence (AI) risk depends on context, impact, and deployment rather than isolated model performance.[8,9] What is missing is a practitioner-level method that captures this contextual layer with benchmark-grade rigor.

This paper presents such a method. TRACE (Tracking Reliability of AI-generated Conversational Evidence) is a practitioner-audit framework for evaluating the downstream workflow reliability of conversational AI. The contribution is the framework, not the 45 incidents. Four instruments operationalize it: a severity rubric grounded in workflow consequence; a workflow-harm taxonomy crosswalked to published taxonomies; a Response-Audit Scorecard that scores the assistant’s behavior after correction; and a User Agency Protocol that converts transient interactions into auditable evidence (Supplementary Appendix S3). The perspective throughout is that of a board-certified clinical informaticist and Certified Professional in Healthcare Quality (CPHQ). The demonstration corpus spans scholarly, clinical informatics, and clinical-adjacent workflows. The framework applies to clinical informatics, quality, and public-health settings, where AI-mediated errors can reach documentation, quality reporting, regulatory artifacts, and care processes. Because the audit records the author’s own de-identified AI-tool outputs rather than identifiable patient data, it demonstrates the framework without making patient-outcome or prevalence claims; those require the multi-site testing outlined in Section 6.

Applied to the demonstration corpus, the framework surfaces a measurement gap we call the claimed-verification gap: the production of verification-implying language without underlying verification. This is the study’s principal empirical finding. It is distinct from confabulation, sycophancy, and over-refusal, and is rarely tested by existing benchmarks. It is best read as a recurring interaction-layer pattern rather than a trait of any one system.

Three research questions organize the paper:

(1) How can practitioner audit logs be structured so that workflow harm, taxonomy mapping, and post-error response become measurable rather than anecdotal?
(2) Which practitioner-observed error categories map cleanly to published taxonomies, and which do not?
(3) What does the demonstration corpus reveal about the claimed-verification gap and about the practical adequacy of post-error response behavior?

## 2. Background and Significance

Three bodies of work most closely surround this study. Each captures something the others miss; none capture the combination this paper offers.

### 2.1 Hallucination, faithfulness, and the benchmark tradition

The benchmark tradition is the most developed. Taxonomies separate factuality from faithfulness failures;[1] benchmarks such as HalluLens evaluate false refusal, hallucination, and false acceptance of nonexistent entities, [10] and cross-lab work shows refusal trades against hallucination exposure.[11] Calibration research finds verbalized confidence can outperform overconfident token probabilities,[5] and human approval of wrong outputs can rise after reinforcement learning from human feedback.[6] Citation fabrication is common in scholarly use,[12] with semantic-entropy detection one route.[13] All of this scores model output, not downstream consequences.

### 2.2 Autoethnography of LLM-assisted scholarship

A growing autoethnographic literature documents researchers’ lived experience with LLMs,[14,15] establishing that researcher experience is legitimate data. Its focus tends toward identity and writing process rather than systematic error analysis with severity coding and taxonomy mapping; it shows practitioners can be the data source, not that they can be the evaluators.

### 2.3 Error-and-harm frameworks in clinical LLM evaluation

The clinical LLM evaluation literature is closest in structure. Asgari et al[16] combined a clinical error taxonomy, a safety framework, and large-scale clinician annotation, reporting low hallucination and omission rates. The difference is the unit of analysis: that work scores LLM-generated clinical notes annotated by clinicians; a practitioner audit scores workflow output judged by the practitioner who bears the consequence.

### 2.4 Human Interaction Evaluation and audit-trail infrastructure

Static benchmarks cannot capture the human-AI interaction surface where many harms occur, motivating Human Interaction Evaluations [17]; process-auditability tools for LLM-assisted qualitative research make a parallel argument.[18] AI incident-reporting and internal algorithmic-auditing frameworks provide infrastructure precedents.[19,20,21] The present method is a low-cost, single-researcher instantiation of that program, anchored in scholarly, clinical informatics, and clinical-adjacent workflows rather than fully autonomous safety-critical systems.

### 2.5 The claimed-verification gap

To our knowledge, none of these literatures directly measures whether a model produces verification-implying language without underlying verification. Hallucination work asks whether the answer is wrong; calibration, whether confidence is appropriate; sycophancy, whether answers shift under pressure. The claimed-verification gap is upstream of all three and recurs across the corpus’s workflows, deserving first-class measurement, with practitioner audit the most tractable route.

Retractions show what happens when flawed content enters the record: they hit record levels in 2023,[22] misconduct drives the majority and error a substantial minority,[23,24] and a recent bibliometric review identifies inappropriate citation and undisclosed machine-generated content as emerging AI-related concerns, noting that plausible but unverified AI content exacerbates peer-review weaknesses.[25] Because retractions occur only after publication, a practitioner audit offers an upstream mechanism for detecting AI-mediated errors before they enter consequential artifacts.

## 3. Materials and Methods

### 3.1 Design

This is a method paper with an empirical demonstration in the reflexive-autoethnographic tradition, reported per the Standards for Reporting Qualitative Research (SRQR) [26]: a longitudinal audit of one researcher’s documented errors over seven weeks, showing what the method produces rather than estimating rates. The audit object is the whole workflow-harm pathway, not the model output alone (Figure 1); the study claims no benchmarking, prevalence estimation, or vendor characterization.

**Figure 1.**
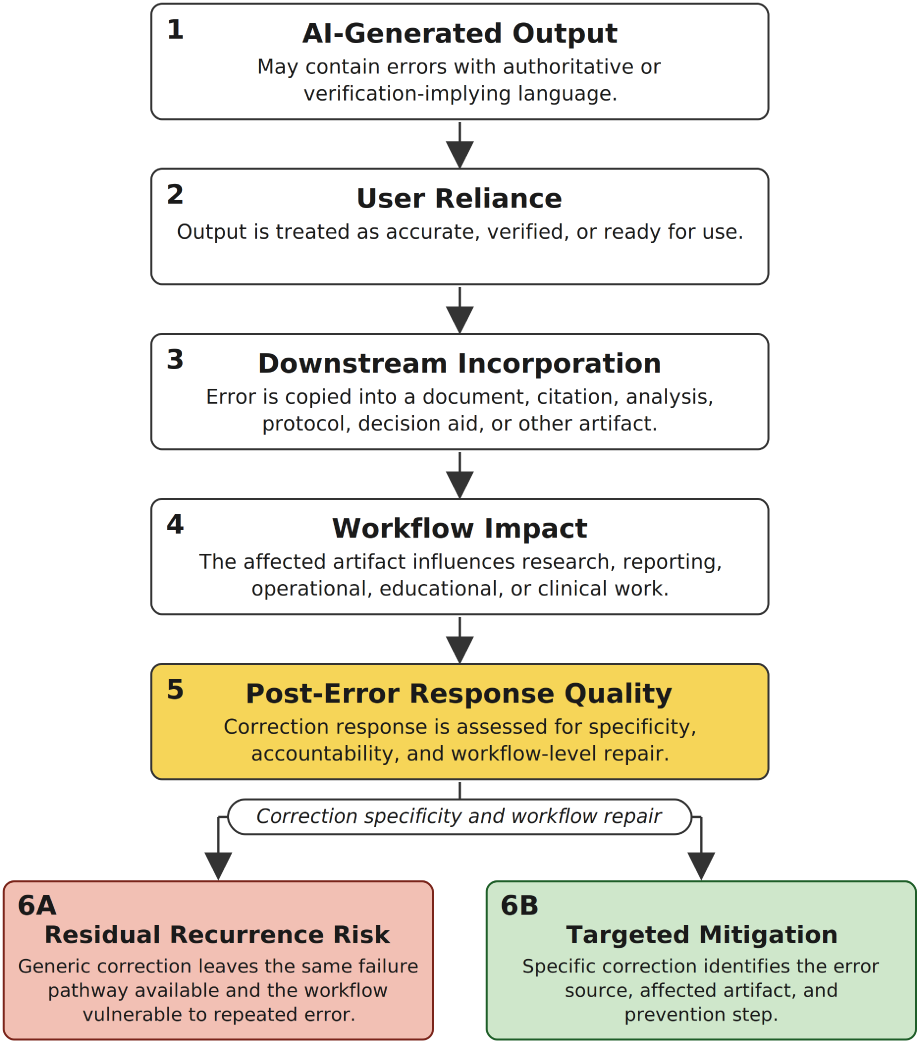
Pathway from AI-generated output to downstream workflow harm. The figure illustrates how an AI-generated output can create workflow harm when erroneous content is presented in authoritative or verification-implying language, treated by users as accurate, and incorporated into downstream artifacts such as documents, citations, analyses, protocols, or decision aids. After an error is identified, the quality of the response determines whether the workflow remains vulnerable to recurrence or whether targeted mitigation addresses the error source, affected artifact, and prevention step. Alt text: Vertical flowchart of six stages. Stage 1, AI-generated output that may contain errors in authoritative or verification-implying language, leads through Stage 2, user reliance; Stage 3, downstream incorporation into a document, citation, analysis, protocol, or decision aid; and Stage 4, workflow impact. Stage 5, post-error response quality, is highlighted as the assessment point and branches, based on correction specificity and workflow repair, to Stage 6A, residual recurrence risk after a generic correction, or Stage 6B, targeted mitigation that addresses the error source, affected artifact, and prevention step.

### 3.2 AI assistance in manuscript preparation: a reflexive disclosure

The practitioner authored the manuscript; the audited system, a frontier commercial conversational LLM, was used as an editing tool under specific revision instructions. It did not originate the conceptual framing, arguments, audit-log content, severity assignments, case selections, or analytic conclusions, and AI-suggested citations were independently retrieved and read before inclusion. Editing-related quality-control issues were tracked separately and informed development of the Response-Audit Scorecard. They are reported in Supplementary Appendix S1 for reflexive transparency and are not part of the primary 45-incident corpus (Supplementary Appendix S2). Consistent with autoethnographic methodology, this reflexive engagement is part of the data but not a substitute for independent evaluation; replication without AI editing assistance is warranted.

### 3.3 Audit corpus

The corpus comprises 45 documented error incidents involving the audited system, recorded by the author from March 15 to May 7, 2026. Workflows spanned scoping-review activities, manuscript development, conference and credentialing materials, regulatory and institutional review board (IRB) drafting, and public-health program review. Model identities were recorded as displayed in the practitioner’s interface, are not independently verified system metadata, and describe the practitioner’s environment rather than a comparison across model versions or vendors.

For rough denominator context, the practitioner’s use of the audited system spanned an estimated several hundred sessions and several thousand exchanges; the 45 documented incidents are the audit-positive subset. The audit is harm-weighted, not prevalence-weighted: it captures incidents consequential enough to document, not the full distribution of output. A de-identified per-incident summary of all 45 incidents is in Supplementary Appendix S2.

#### 3.3.1 Documentation threshold and the audit-positive subset

The 45 incidents are documentation-positive errors only and under-represent the true error rate: the threshold was consequence-driven, so errors caught at generation, judged too local, or resolved inline went unrecorded. Appropriate for a workflow-level audit, this means the severity distribution (9 critical, 12 high, 18 medium, 6 low) overrepresents severe incidents; the implied per-session rate is a lower bound; and 45 is not a frequency estimate.

Adequacy rests on saturation rather than enumeration: recurring patterns and the claimed-verification finding stabilized within the documented set, and a future session-reconstruction density check is recommended (Section 6).

### 3.4 Definition of an error

An incident entered the log when an output or interaction met at least one of seven criteria: factual inaccuracy in a numerical, regulatory, bibliographic, or domain claim; unsupported claim presented as fact or with implied source support; false or implied verification, where the model behaved as though it had checked a source, tool, file, or prior context when it had not. The remainder: context or workflow contradiction, including loss of prior task constraints; tool or system misunderstanding; document-integrity failure; and refusal or response-pattern inconsistency relevant to workflow reliability.

The definition deliberately includes both output-level and interaction-level failures. In real workflows, harm can arise from the content of a wrong answer, the way that answer is packaged, or the assistant’s behavior after correction.

### 3.5 Verification regime as a methodological condition

The audit ran under a layered verification regime shaped by the author’s healthcare-quality background (CPHQ) and Reason’s Swiss-cheese model [27,28]. Claims involving regulation, statistics, citations, eligibility, tool capabilities, or procedures were treated as unverified until corroborated, with depth scaled to consequence. The 45 incidents are thus errors that remained visible despite this posture, not the full error population; different regimes would yield different audit-positive subsets (Section 6).

### 3.6 Severity rubric

Severity was coded by consequence, not by embarrassment, annoyance, or model confidence (Table 1):

**Table 1.**
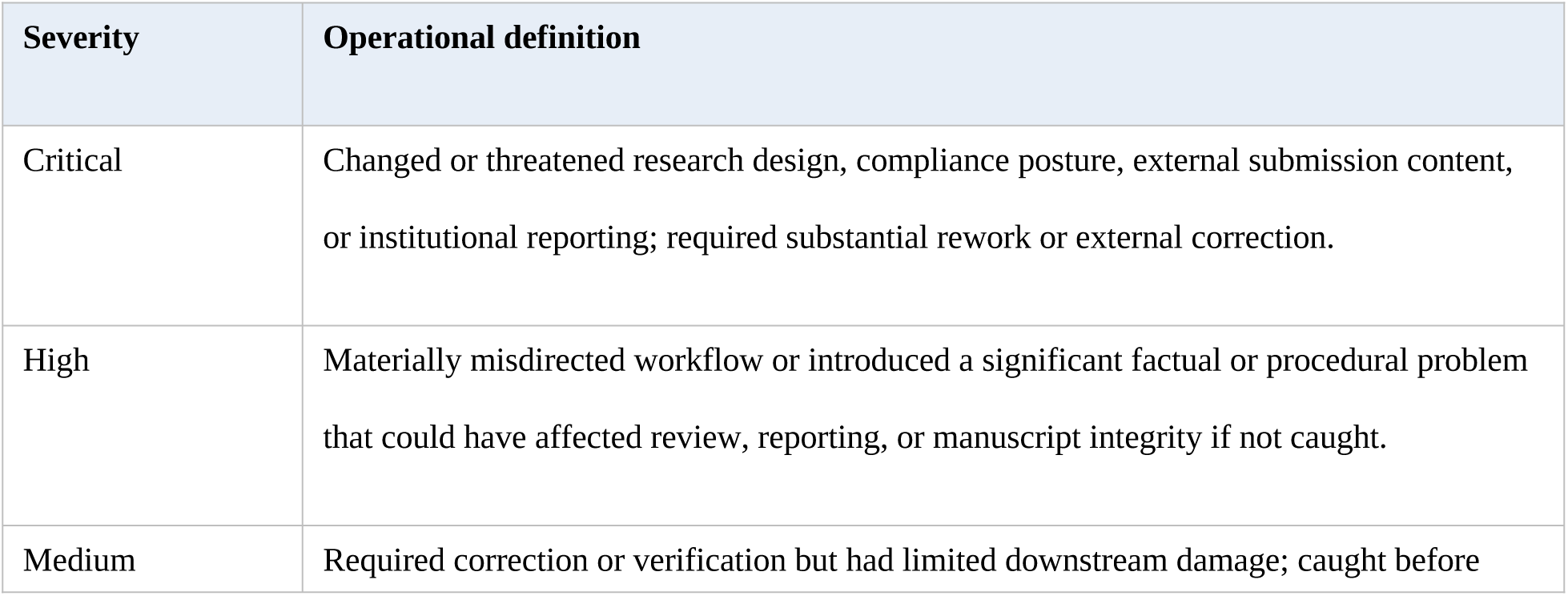
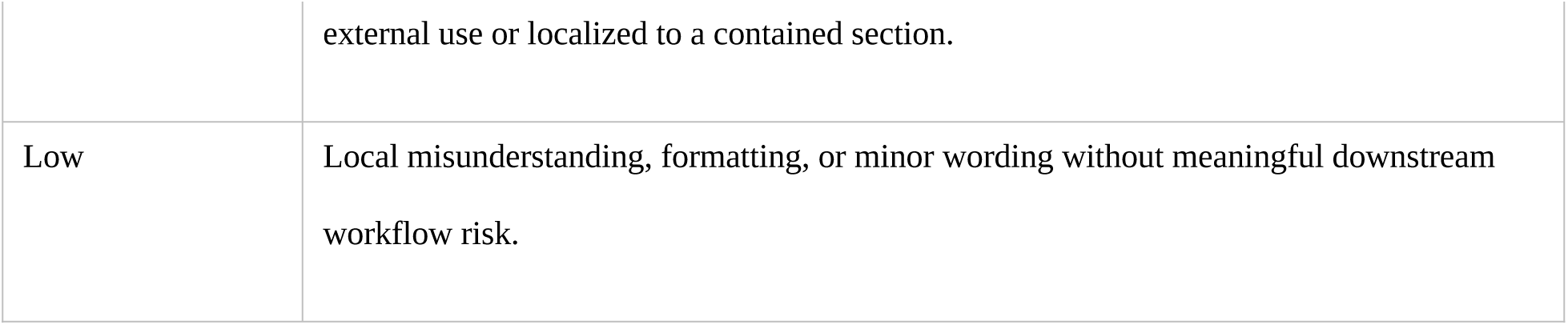
Consequence-based severity rubric. The resulting distribution was 9 critical, 12 high, 18 medium, and 6 low. These are incident counts, not rates.

| Severity | Operational definition |
| --- | --- |
| Critical | Changed or threatened research design, compliance posture, external submission content, or institutional reporting; required substantial rework or external correction. |
| High | Materially misdirected workflow or introduced a significant factual or procedural problem that could have affected review, reporting, or manuscript integrity if not caught. |
| Medium | Required correction or verification but had limited downstream damage; caught before |
|  | external use or localized to a contained section. |
| Low | Local misunderstanding, formatting, or minor wording without meaningful downstream workflow risk. |

### 3.7 Coding procedure and intercoder considerations

Primary coding was performed by the practitioner-author, who normalized entries into recurring categories; because the author was both practitioner and analyst, this is first-person practitioner-audit evidence rather than blinded adjudication.

Three authors (M.A., B.M., J.E.L.) independently coded a stratified random subsample of 16 incidents (4 critical, 5 high, 5 medium, 2 low; Python Mersenne Twister, seed 20260509). Each was blinded to the practitioner-author’s coding and to the others, and assigned a category, severity, and a claimed-verification flag (Pack 1). Each then received a second pack (Pack 2) displaying the practitioner-author’s coding for the same incidents and recorded agreement judgments. Distribution logistics, a documented sequential-gating deviation for one reviewer-author, and signed blinding certifications are recorded in the audit trail. Agreement used Fleiss’ kappa for nominal dimensions, pairwise Cohen’s kappa, and quadratic-weighted kappa for ordinal severity (Table 2).

**Table 2.**
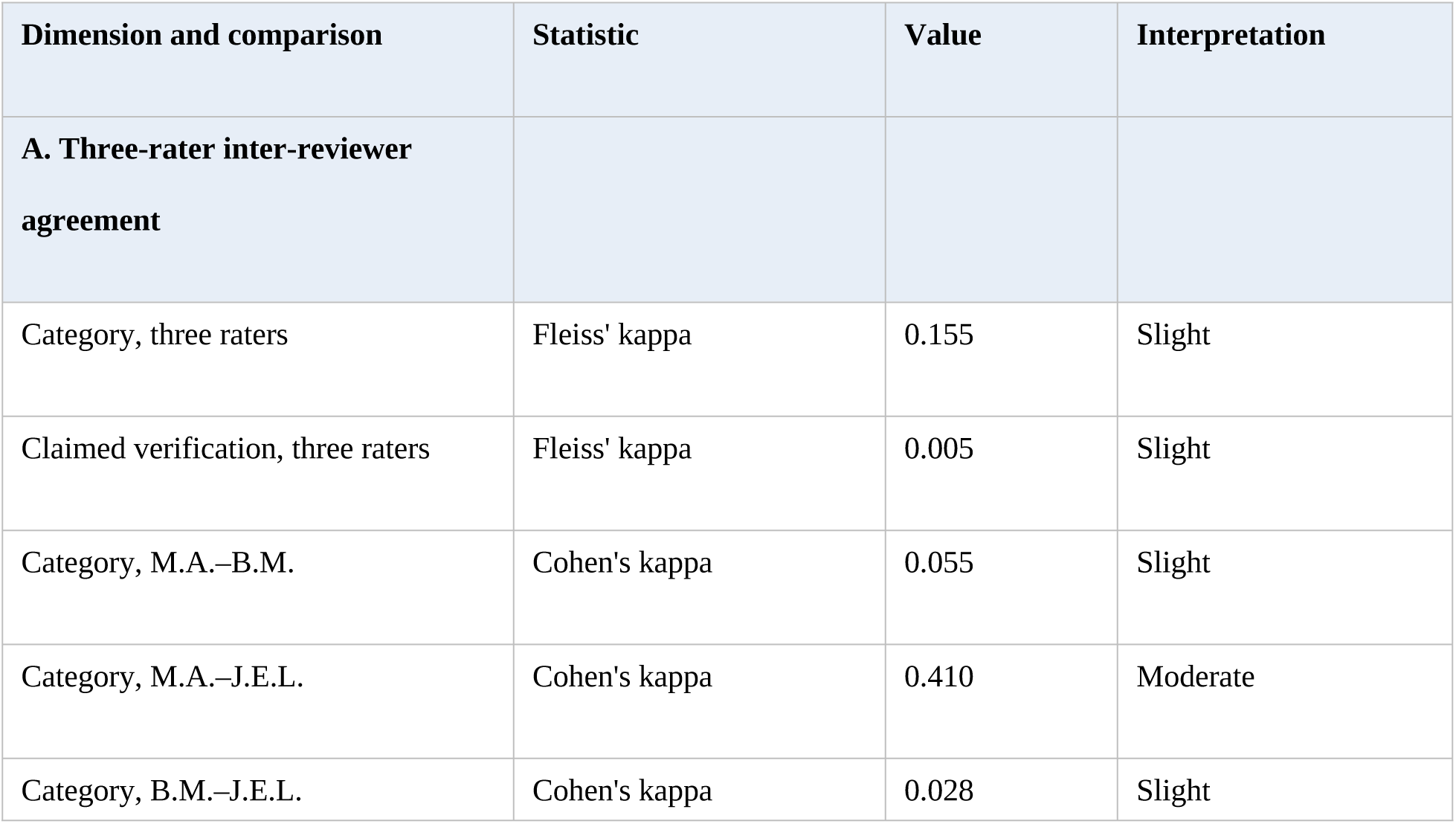
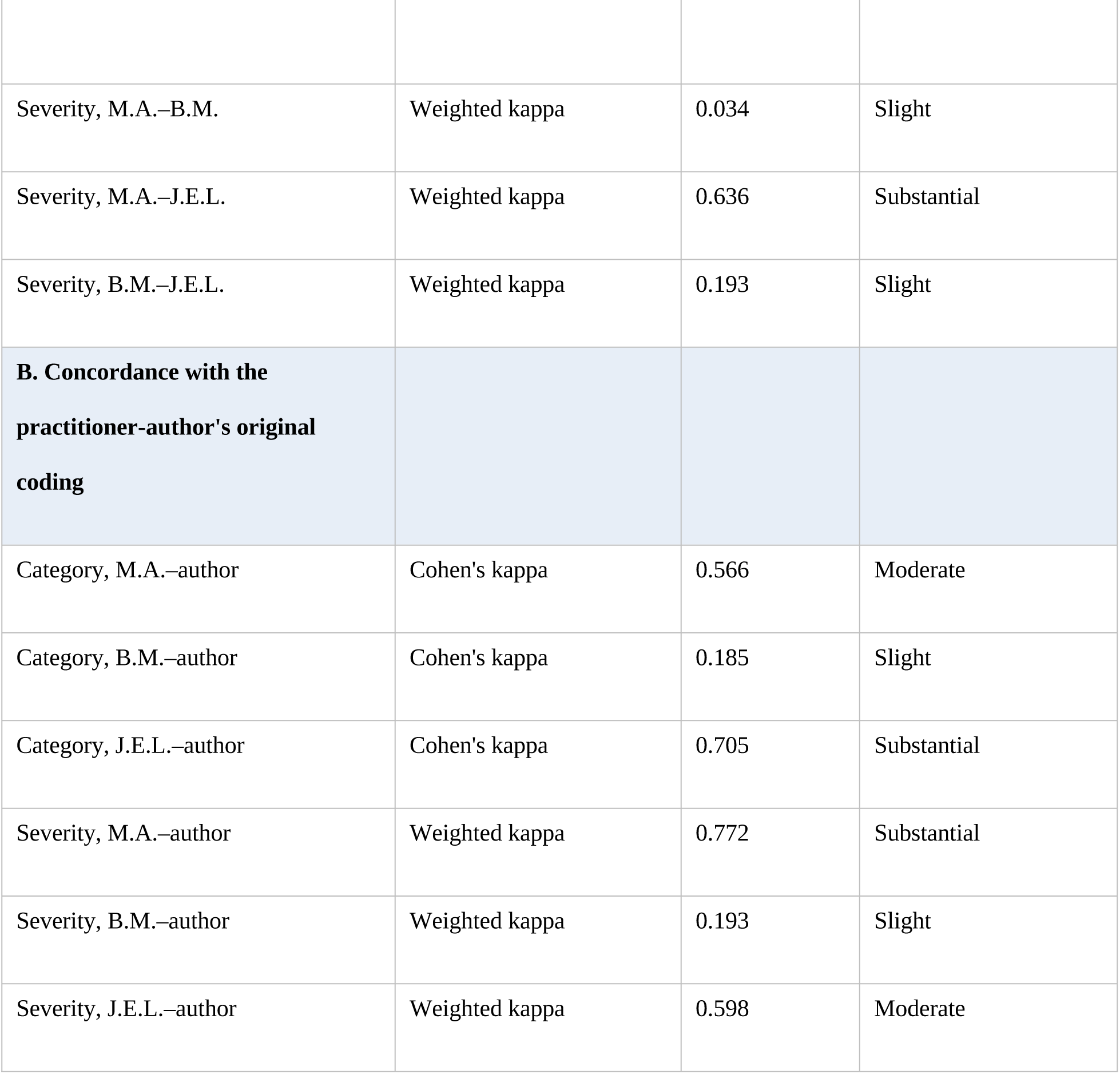
Inter-rater reliability on the 16-incident stratified subsample (Pack 1). Panel A: agreement across the three reviewer-authors. Panel B: each reviewer-author’s concordance with the practitioner-author’s original coding. Pack 2 descriptive agreement with the practitioner-author—category: M.A. 10/16, B.M. 5/16, J.E.L. 12/16; severity judged appropriate: M.A. 11/16, B.M. 6/16, J.E.L. 4/16.

Inter-reviewer agreement was low (Table 2): three-rater category agreement reached Fleiss’ kappa 0.155, and the claimed-verification flag only 0.005.[29] Disagreement was structured rather than random: M.A. and J.E.L. converged moderately (category 0.410; severity 0.636), whereas B.M. diverged from both (0.055 and 0.028). Concordance with the practitioner-author’s original coding (Table 2, panel B) was highest for J.E.L. and lowest for B.M.; these values describe concordance with the original practitioner coding and are not independent reliability estimates. All three reviewer-authors independently identified overlapping categories, missing codes, and an under-specified claimed-verification dimension, and all three generally assigned lower severity than the practitioner-author, a pattern consistent with severity judgments being sensitive to whether the coder directly experienced the downstream consequence. This asymmetry may reflect practitioner proximity to consequence, practitioner overestimation, or both; the present design cannot distinguish these explanations. Each reviewer-author’s Pack 2 judgments were fully consistent with their own Pack 1 codes, indicating the two-pack design produced coherent comparison rather than post hoc endorsement. The low agreement is interpreted (Sections 5.3, 7) as a finding about the difficulty of taxonomizing LLM-mediated workflow errors, not a refutation of the corpus.

### 3.8 The Response-Audit Scorecard

The Response-Audit Scorecard (Table 3) scores post-error behavior on seven criteria (specific acknowledgment, failure-mode explanation, evidence correction, harm recognition, prevention mechanism, transparency limits, recurrence tracking). It was developed iteratively in response to post-error replies that offered apology and generic reassurance without naming the failure or a safeguard. It makes correction behavior part of the audit object: two assistants that produce the same error but differ in post-error response have different reliability profiles.

**Table 3.**
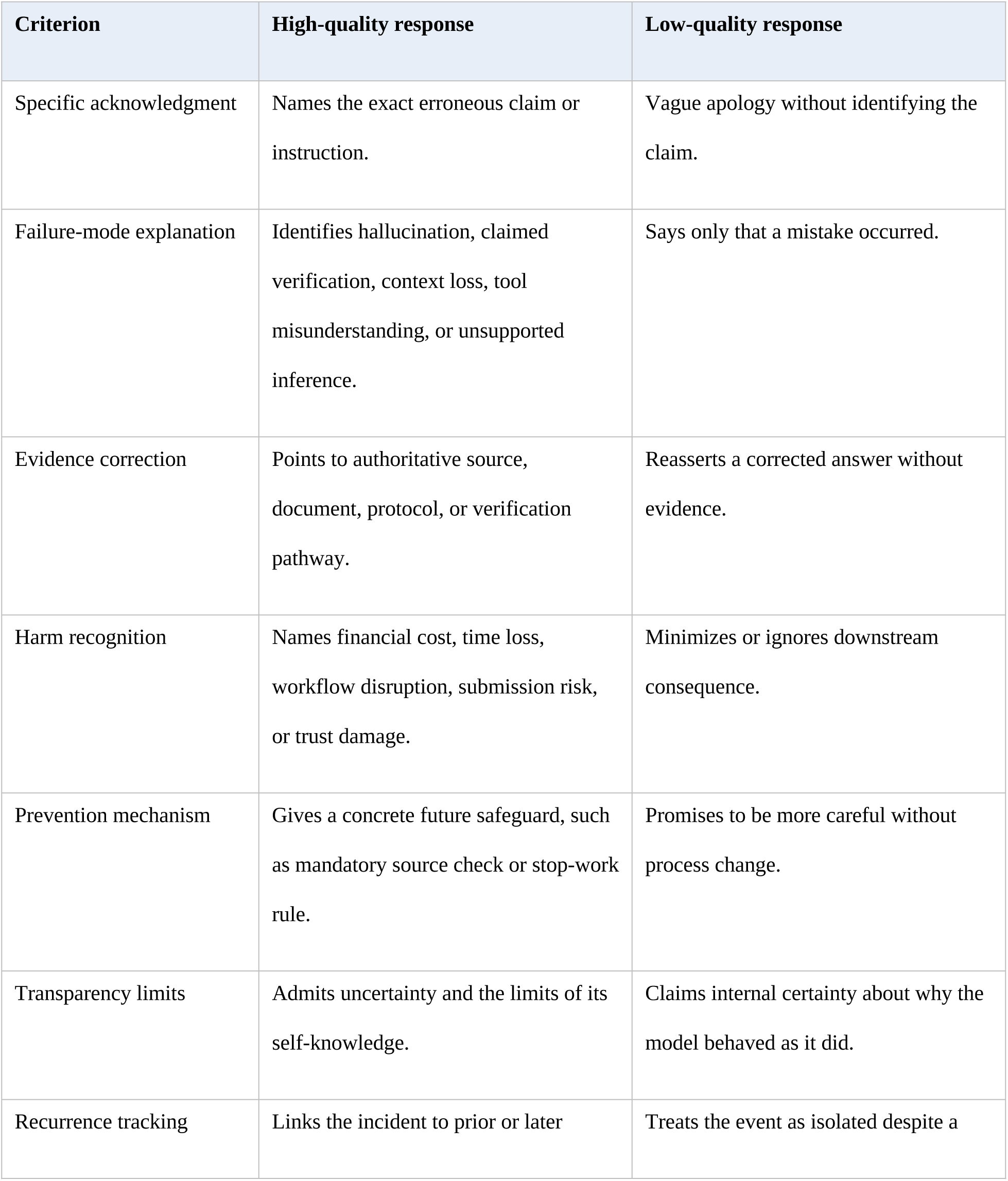
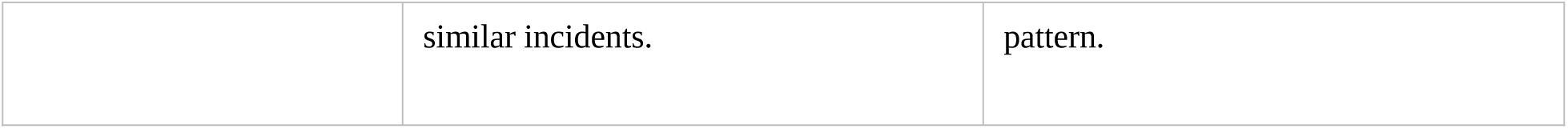
Response-Audit Scorecard: high-versus low-quality post-error response criteria.

### 3.9 Taxonomy mapping procedure

Practitioner categories were mapped to published failure categories in three steps: identify the model behavior at the output or interaction level; identify the downstream workflow consequence; and map the practitioner code to one or more published categories (factuality hallucination, faithfulness failure, context inconsistency, calibration failure, false refusal, sycophancy, citation fabrication, or claimed verification). The mapping is interpretive, positioning incidents within the evaluation literature without formal benchmarking.

### 3.10 Comparative literature selection

The comparative literature was purposive, not systematic, selected to represent major LLM failure families relevant to the corpus, not intended to be exhaustive.

### 3.11 Ethics, IRB exemption rationale, and Terms of Service

This study did not involve human-subjects research as defined by 45 CFR 46.102(e)(1): the data are the author’s own AI tool outputs, with no other individuals studied and no identifiable third-party information. The Xavier University of Louisiana Institutional Review Board determined on July 6, 2026 that the study does not constitute human subjects research and does not require IRB approval; the project was not assigned a study number. Incident examples are de-identified; no patient-, student-, reviewer-, or collaborator-identifiable data or proprietary system metadata are included. The author verified applicable platform terms.

Figure 2 shows how the four operational instruments of the TRACE practitioner-audit method map onto the workflow-harm pathway stages introduced in Figure 1. The Workflow-Harm Taxonomy classifies incidents at the model-output, user-reliance, and downstream-incorporation stages. The Severity Rubric assigns consequence-based severity at the workflow-impact stage. The Response-Audit Scorecard (Table 3) evaluates post-error assistant behavior at the evaluation point (Stage 5). The User Agency Protocol (Supplementary Appendix S3) spans all pathway stages as the practitioner’s active response infrastructure.

**Figure 2.**
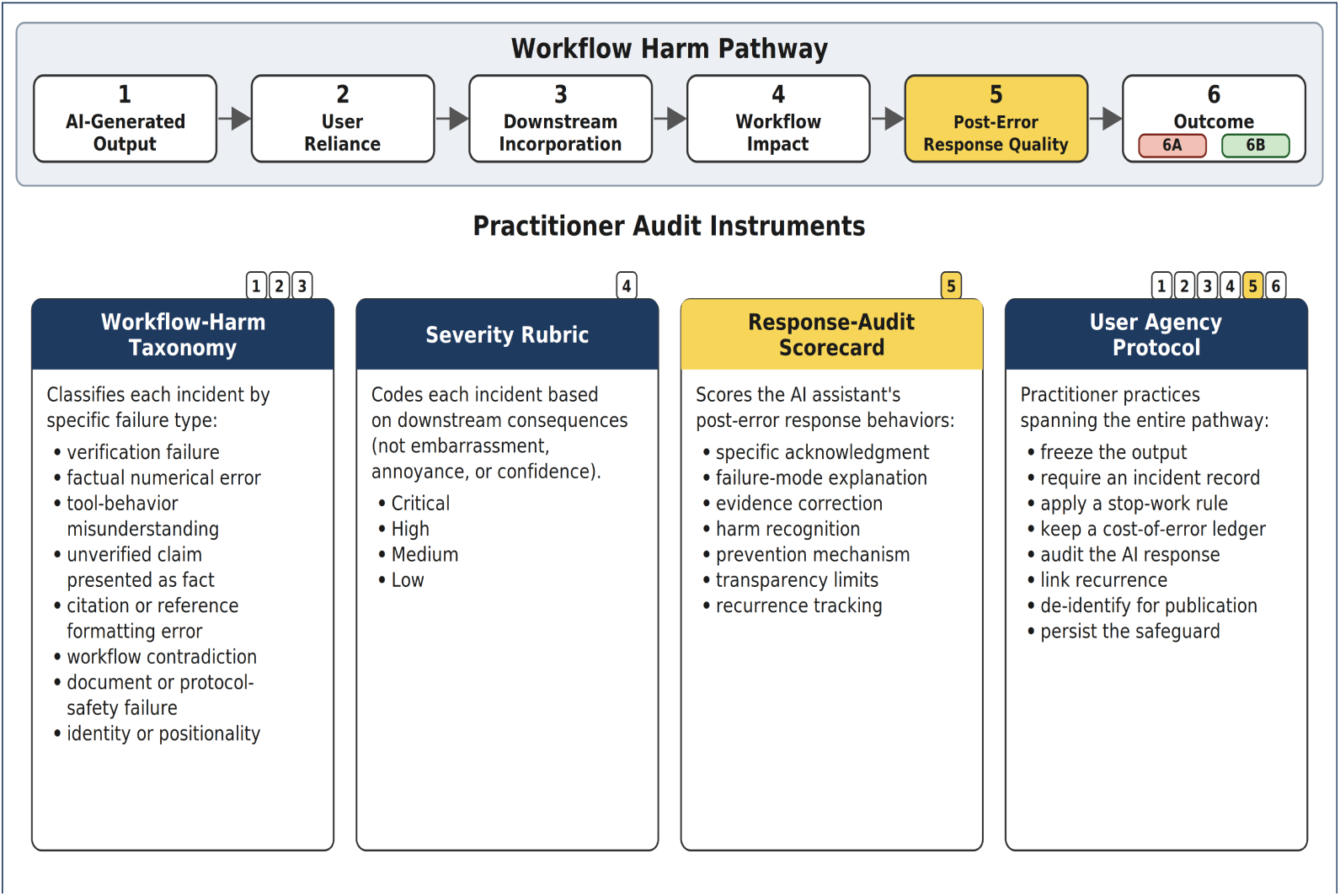
TRACE practitioner-audit method: tracking reliability of AI-generated conversational evidence. Six workflow-harm pathway stages (top row) and four practitioner-audit instruments (bottom row). Stage 5 (Post-Error Response Quality) is highlighted as the primary evaluation point where the Response-Audit Scorecard applies. Outcomes 6A (Residual Recurrence Risk) and 6B (Targeted Mitigation) branch from Stage 5. Alt text: Diagram of the TRACE practitioner-audit method. A top row shows the six workflow-harm pathway stages, with Stage 5, post-error response quality, highlighted and Stage 6 split into outcomes 6A and 6B. Below, four instrument panels—Workflow-Harm Taxonomy, Severity Rubric, Response-Audit Scorecard, and User Agency Protocol—list the eight locked failure categories, the four severity levels, the seven post-error response criteria, and the eight practitioner operating practices, each panel tagged with the pathway stages to which it applies.

### 3.12 Supplemental AI-comparator analysis of taxonomy interpretability

As a supplemental, exploratory extension, three independent frontier LLM comparators were prompted to apply the locked eight-category taxonomy to all 45 corpus incidents. The code set was locked before reviewer coding and this comparator analysis; the practitioner-author’s full-corpus incident assignments were finalized under the written coding conventions, with category definitions and the 13-to-8-category crosswalk, in Supplementary Appendix S2. Comparators were prompted under standardized, blinded instructions at temperature 0. Each independently coded category, severity, and claimed-verification flag from the same de-identified descriptions used in the human coding packs. Cross-system agreement was computed with Fleiss’s kappa (three raters) and pairwise Cohen’s kappa. This analysis assesses whether independent systems apply the category scheme consistently (legibility); it is not a reliability statistic, a validation of the incidents, or a substitute for human coding. The audited system was excluded from the comparator set to avoid circularity and is not named, to avoid converting a method paper into a product evaluation; comparator identities are reported collectively, with exact model identifiers and run parameters retained in the audit trail for editor review. A 10-category sensitivity analysis added the two reviewer-motivated categories (Section 5.3); materials are in Supplementary Appendix S5.

## 4. Results

### 4.1 Corpus summary

Across seven weeks, 45 error incidents involving the audited system were documented. The severity distribution was 9 critical, 12 high, 18 medium, and 6 low. The four most frequent normalized categories tied: verification failure, factual numerical error, tool-behavior misunderstanding, and citation or reference formatting error (n=7 each), followed by document or protocol-safety failure (n=6), workflow contradiction and unverified claim presented as fact (n=4 each), and identity or positionality (n=3). Per-incident coding, the 13-to-8-category crosswalk, and coding conventions appear in Supplementary Appendix S2. The most consequential incidents were not confined to any one category: workflow contradiction and protocol-safety failure carried the highest downstream risk, affecting research procedure, screening integrity, or submission readiness. Supplementary Appendix S2 provides a finer-grained 13-category normalization of the same 45 incidents; the two taxonomies are parallel views, not a counting discrepancy.

### 4.2 De-identified incident examples

These illustrative incidents (Table 4) show why severity cannot be inferred from output type alone: a hallucinated number may be low-consequence in brainstorming and high-consequence in institutional reporting, and a context inconsistency minor in casual writing but critical in a multi-step research workflow.

**Table 4.** De-identified incident examples with severity and taxonomy mapping. Cases 2 and 5 are composite examples synthesizing recurring incident patterns; their severity entries show the range recorded across the corresponding corpus incidents (Supplementary Appendix S2). Single-severity rows reflect individually recorded incidents.

| Case | Workflow | Model behavior | Severity | Taxonomy mapping |
| --- | --- | --- | --- | --- |
| 1 | Scoping review | Contradictory procedural guidance about reviewer settings and screening workflow. | Critical | Context inconsistency; workflow safety failure; procedural hallucination. |
| 2 | Manuscript development | Truncation or corruption of manuscript material during editing or revision. | High–Critical | Context loss; document-integrity failure; faithfulness failure. |
| 3 | Program review | Fabricated enrollment value presented with apparent confidence. | High | Factual fabrication; numerical hallucination; verification failure. |
| 4 | IRB-adjacent drafting | Unsupported regulatory claim presented as usable guidance. | High | Domain-specific factuality failure; overconfidence; claimed verification. |
| 5 | Error-response audit | Post-error explanation acknowledged patterns but | Medium–Critical | Response-audit failure; correction-behavior |
|  |  | risked generic reassurance and unkept commitments. |  | calibration; interaction-layer reliability. |

### 4.3 The claimed-verification finding

Among the most frequent normalized categories was verification failure: procedural language implying that a source, tool state, prior context, or computation had been checked when it had not—regulatory citations rendered confidently, statistics described as cross-checked, prior-chat content phrased as recall. Distinct from confabulation, sycophancy, and over-refusal, its shared feature is verification-implying language without underlying verification.

It is a recurring interaction-layer pattern: a model trained on careful scholarly prose reproduces the verbal markers of having checked regardless of whether retrieval or computation occurred, and a practitioner can detect the gap by verifying against authoritative sources what the assistant cannot.

### 4.4 Workflow-harm patterns

Four workflow-harm patterns recurred. Procedural propagation: an output shaped how a workflow was executed, with errors compounding across steps. Documentary contamination: erroneous content entered a manuscript, citation list, report, or submission—an integrity-level rather than formatting-level harm. Trust-calibration disruption: confident tone or claimed verification degraded the user’s ability to allocate trust. User-borne corrective burden: the error imposed direct corrective costs, added software needs, and unrecoverable labor.

### 4.5 Direct-cost case with counterfactual

One scoping-review incident carried a practitioner-estimated corrective impact of approximately $2500; a practitioner-estimated composite of reviewer hours, screening-restart costs, and third-reviewer adjudication (Supplementary Appendix S2), not a formal cost analysis. Beyond direct costs, the incident delayed scheduled funder deliverables, website development, and an institutional review board submission. Contradictory reviewer-settings guidance, acted on mid-screening, irreversibly locked 3,130 records under single-reviewer mode and reduced the planned dual-review overlap from 4,166 to 1,164 records; the resulting κ = 0.224 on the residual overlap breached the prespecified κ ≥ 0.70, forcing a screening restart on a separate platform. This screening-phase statistic is a consequence within the audited incident, not a property of this study’s coding (Section 3.7).

A conservative reading treats the AI contribution as necessary but not sufficient, consistent with research-waste[30,31] and screening-workload[32,33] literatures; detailed entries are in Supplementary Appendix S2.

### 4.6 Response-audit findings

Applying the Response-Audit Scorecard to post-error interactions showed variable correction quality. Higher-quality responses named the error and failure mode, cited an authoritative source, acknowledged the consequence, and proposed a safeguard; lower-quality responses offered generic apology or restated a corrected answer without evidence.

The scorecard exposes recurrence risk that conventional metrics miss; the present corpus motivates, but is too small to demonstrate, this quantitatively.

### 4.7 Editing quality-control observations

The AI-assisted editing process generated quality-control observations involving citation, attribution, regulatory language, and calibration issues. These observations informed the AI Editing Disclosure and manuscript verification process but are not included in the primary 45-incident empirical corpus and are not used to estimate error frequency. Across the audit corpus, no documented mistake was flagged spontaneously by the audited system.

### 4.8 Supplemental AI-comparator interpretability

In the supplemental AI-comparator analysis, category-level agreement across the three comparators was substantial (Fleiss κ = 0.632 across all 45 incidents), whereas agreement for severity (κ = 0.401) and the claimed-verification flag (κ = 0.405) was fair. Comparator agreement was higher than the human category agreement reported in Section 3.7 (Fleiss κ = 0.155); this reflects taxonomy legibility under standardized, fixed-instruction conditions rather than superior comparator judgment: the comparators faced none of the interpretive variability human raters bring. Severity and claimed verification stayed judgment-dependent. A standalone 10-category sensitivity analysis did not reduce category-level interpretability. These results are exploratory and hypothesis-generating and signal taxonomy legibility rather than independent validation. Because the comparators may share training data and architecture, their agreement may partly reflect shared model priors; the analysis is best read as triangulation against, not confirmation of, the human coding. Full materials are in Supplementary Appendix S5.

## 5. Discussion

The manuscript does not argue that conversational LLMs lack utility; the audited system was itself the editing tool here. The argument is that their usefulness in clinical and clinical-adjacent workflows requires structured verification proportional to consequence, which current evaluation approaches have not operationalized.

### 5.1 What benchmarks measure well

Formal evaluation supplies useful vocabulary: the hallucination, long-context, calibration, and sycophancy literatures reviewed in Section 2.1 name most of what went wrong at the model-output level.

### 5.2 What practitioner audits add

The framework captures what benchmarks do not: downstream consequence, longitudinal recurrence across heterogeneous tasks, post-error response behavior, and the claimed-verification gap. It complements formal evaluation rather than replacing it. Across the corpus, the audited system never spontaneously flagged an error. Every correction was human-initiated (Supplementary Appendix S1), underscoring that verification in these workflows remained a practitioner responsibility.

### 5.3 The claimed-verification gap as a research agenda

The claimed-verification gap should be evaluated alongside hallucination, and remains traceable in real workflows even where it resists operationalization in static benchmarks. Benchmarks could label verification-implying language and check whether implied verification corresponds to retrieval or tool use; interfaces could expose verification state; and evaluation suites could test resistance to producing verification-implying language absent retrieval.

All three reviewer-authors independently converged on taxonomy refinements—an identifier-error code, a behavioral/interactional-pattern code, a more concrete claimed-verification definition, and a discrete-versus-aggregated consequence distinction—strengthening the argument that these errors resist clean taxonomization. The supplemental AI-comparator analysis (Sections 3.12, 4.8) is consistent with this: adding the two reviewer-motivated categories did not reduce category-level interpretability.

### 5.4 Implications for assistant design and governance

The findings support a conservative rule for high-stakes AI assistance: treat every model claim involving regulation, statistics, citations, eligibility, tool capabilities, or procedures as unverified until checked, consistent with vendor system-card cautions[2,3] and resilience frameworks emphasizing ongoing monitoring.[34] The conclusion is not that such systems should never be used, but that high-impact use requires structured verification, stop-work rules, and audit trails proportional to consequence.

### 5.5 Single-practitioner design and generalizability

TRACE is a framework, not a characterization of any vendor or a definitive error rate. The single-practitioner, single-system design provides longitudinal depth but not prevalence or cross-model claims (Section 7); the multi-site and multi-model work in Section 6 is required first.

## 6. Future Directions

This is a foundational pilot. Priority work includes extending multi-coder reliability testing to the full corpus and multi-site replication across practitioners and workflows. It also includes multi-model replication testing whether the claimed-verification gap recurs across structurally different LLMs; a session-reconstruction density check; longitudinal testing of whether Response-Audit Scorecard ratings predict recurrence; and integration with institutional safety-event and quality-improvement infrastructure. None requires vendor-internal access; the framework is executable with user-facing interfaces and de-identified logs. A step-by-step replication workflow is provided in Supplementary Appendix S4.

## 7. Limitations

Several limitations bear on interpretation. The design is observational and single-practitioner, conducted in one institutional context, so the findings describe a framework in use rather than a population. The corpus contains documentation-positive incidents only (Section 3.3.1); the count is not a frequency estimate, and the severity distribution is not representative. Coding was retrospective. The taxonomy is still evolving: all three reviewer-authors identified overlapping categories, missing codes, and an under-specified claimed-verification dimension (Section 3.7). Because the practitioner-author recorded, coded, and analyzed his own incidents, confirmation bias cannot be excluded; the independent three-reviewer check, the audit trail, and the AI Editing Disclosure mitigate but do not eliminate it. The framework also depends on practitioner expertise: recognizing an incident presupposes the domain knowledge to notice that an output is wrong, so audit yield varies with the auditor. The three-reviewer check showed low agreement, attributed to coding-scheme limitations and rater heterogeneity rather than instability of the incident-level signal; one reviewer-author received the comparison pack before returning the blinded pack (a documented deviation with signed certifications). The comparative literature was purposive, not systematic. Model identity was taken as recorded, without independent metadata, and should not be read as a vendor comparison. The study evaluates interaction-layer workflow phenomena rather than model architecture or capability under controlled conditions.

## 8. Conclusion

Real-world LLM reliability is not adequately described by output correctness alone. In clinical and clinical-adjacent workflows, model output becomes a workflow-level reliability concern when fluent, confident, partially wrong content reaches procedures, documents, citations, reporting, or regulatory language. The contribution of this paper is TRACE: a practitioner-audit framework for evaluating the downstream workflow reliability of conversational AI. It pairs model output with task context, downstream consequence, and correction behavior. Applied to the demonstration corpus, the framework surfaces the claimed-verification gap, a recurring interaction-layer pattern that benchmarks rarely test directly. The pattern warrants testing across conversational AI systems rather than being assumed to be vendor-specific. Formal benchmarks remain essential; the framework complements them by showing how model behavior affects real work and how assistants behave when corrected, especially where errors are consequential.

## Data Availability

The de-identified audit corpus underlying this study, including raw and normalized category labels, severity ratings, error summaries, correction sources, and downstream consequences for all 45 incidents, is provided in Supplementary Appendix S2. Additional anonymized excerpts are available from the corresponding author on reasonable request, subject to the redaction protocol described in the manuscript. No proprietary system metadata, model weights, or third-party identifiable data are included. The full audit-log coding schema (the per-incident data structure) and the User Agency Protocol are provided in Supplementary Appendix S3.

## AI Editing Disclosure

The authors declare the use of generative AI in the research and writing process. According to the GAIDeT taxonomy[35] (2025), the following tasks were delegated to GAI tools under full human supervision:

- Literature search and systematization
- Visualization
- Proofreading and editing
- Reformatting
- Publication support
- Supplemental AI-comparator taxonomy-interpretability coding (Section 3.12), under full human supervision

Generative AI tools were used in three roles: manuscript-support tools (drafting, editing, formatting, and literature systematization under author direction); AI comparators for the supplemental taxonomy-interpretability analysis (Section 3.12); and the audited conversational system that is the subject of the empirical corpus. The audited system is vendor-blinded in the manuscript because the study evaluates interaction-layer workflow phenomena rather than vendor-specific performance. Full tool identities and exact model identifiers for all three roles are retained in the study audit trail and can be provided to editors on request.

Responsibility for the final manuscript lies entirely with the authors. GAI tools are not listed as authors and do not bear responsibility for the final outcomes. Declaration submitted by Davis Austria.

## Author Contributions (Contributor Roles Taxonomy [CRediT])

D.A.: Conceptualization, Methodology, Investigation, Data Curation, Formal Analysis, Visualization, Project Administration, Writing – Original Draft, Writing – Review & Editing. B.M.: Investigation, Formal Analysis, Validation, Writing – Review & Editing. J.E.L.: Investigation, Formal Analysis, Validation, Writing – Review & Editing. M.A.: Methodology, Investigation, Validation, Writing – Review & Editing. M.O.: Supervision, Writing – Review & Editing. All authors approved the final manuscript.

## Supporting information

Supplementary Appendix S1: AI-Assisted Editing Quality-Control Log and Redaction Protocol

Supplementary Appendix S2: De-identified Audit Corpus Summary

Supplementary Appendix S3: Audit-Log Coding Schema and User Agency Protocol

Supplementary Appendix S4: Replication Workflow for Practitioner Audits

Supplementary Appendix S5: AI-Comparator Analysis Materials and Results

## References

1. Huang L, Yu W, Ma W, et al. A survey on hallucination in large language models: principles, taxonomy, challenges, and open questions. arXiv. Preprint posted online November 2023. arXiv:2311.05232

2. Anthropic. The Claude 3 Model Family: Opus, Sonnet, Haiku. 2024. Accessed May 9, 2026. https://assets.anthropic.com/m/61e7d27f8c8f5919/original/Claude-3-Model-Card.pdf

3. OpenAI. GPT-4 system card. March 2023. Accessed May 9, 2026. https://cdn.openai.com/papers/gpt-4-system-card.pdf

4. Liu NF, Lin K, Hewitt J, et al. Lost in the middle: how language models use long contexts. Trans Assoc Comput Linguist. 2024;12:157–173.

5. Tian K, Mitchell E, Yao H, Manning CD, Finn C. Fine-tuning language models for factuality. In: Proceedings of the Twelfth International Conference on Learning Representations (ICLR); 2024. arXiv:2311.08401

6. Wen J, Zhong R, Khan A, et al. Language models learn to mislead humans via RLHF. arXiv. Preprint posted online September 19, 2024. arXiv:2409.12822

7. Fanous A, Goldberg J, Agarwal AA, et al. SycEval: evaluating LLM sycophancy. arXiv. Preprint posted online February 12, 2025. arXiv:2502.08177

8. National Institute of Standards and Technology. Artificial Intelligence Risk Management Framework (AI RMF 1.0). NIST AI 100-1. January 2023. doi:10.6028/NIST.AI.100-1

9. National Institute of Standards and Technology. Artificial Intelligence Risk Management Framework: Generative Artificial Intelligence Profile. NIST AI 600-1. July 2024. doi:10.6028/NIST.AI.600-1

10. Bang Y, Ji Z, Schelten A, et al. HalluLens: LLM hallucination benchmark. In: Proceedings of the 63rd Annual Meeting of the Association for Computational Linguistics (Volume 1: Long Papers). Association for Computational Linguistics; 2025:24128–24156.

11. OpenAI, Anthropic. OpenAI-Anthropic safety evaluation. 2025. Accessed May 9, 2026. https://openai.com/index/openai-anthropic-safety-evaluation/

12. Linardon J, Jarman HK, McClure Z, Anderson C, Liu C, Messer M. Influence of topic familiarity and prompt specificity on citation fabrication in mental health research using large language models: experimental study. JMIR Ment Health. 2025;12:e80371. doi:10.2196/80371

13. Farquhar S, Kossen J, Kuhn L, Gal Y. Detecting hallucinations in large language models using semantic entropy. Nature. 2024;630(8017):625–630. doi:10.1038/s41586-024-07421-0

14. Schwenke N, Söbke H, Kraft E. Potentials and challenges of chatbot-supported thesis writing: an autoethnography. Trends High Educ. 2023;2(4):611–635. doi:10.3390/higheredu2040037

15. Yang S, Liu Y, Wu T-C. ChatGPT, a new “ghostwriter”: a teacher-and-students poetic autoethnography from an EMI academic writing class. Digit Appl Linguist. 2024;1:2244. doi:10.29140/dal.v1.2244

16. Asgari E, Montaña-Brown N, Dubois M, et al. A framework to assess clinical safety and hallucination rates of LLMs for medical text summarisation. NPJ Digit Med. 2025;8:274. doi:10.1038/s41746-025-01670-7

17. Ibrahim L, Huang S, Ahmad L, Anderljung M. Beyond static AI evaluations: advancing human interaction evaluations for LLM harms and risks. arXiv. Preprint posted online May 2024. arXiv:2405.10632

18. Lu MH, Ellegood R, Rodriguez-Ramirez R, Blumert S. Affording process auditability with QualAnalyzer: an atomistic LLM analysis tool for qualitative research. arXiv. Preprint posted online April 2026. arXiv:2604.03820

19. Raji ID, Smart A, White RN, et al. Closing the AI accountability gap: defining an end-to-end framework for internal algorithmic auditing. In: Proceedings of the 2020 Conference on Fairness, Accountability, and Transparency (FAT* 2020). Association for Computing Machinery; 2020:33–44. doi:10.1145/3351095.3372873

20. McGregor S, Ettinger A, Judd N, et al. To err is AI: a case study informing LLM flaw reporting practices. arXiv. Preprint posted online October 15, 2024. arXiv:2410.12104

21. Paeth K, Atherton DS, Pittaras N, Frase H, McGregor S. Lessons for editors of AI incidents from the AI Incident Database. arXiv. Preprint posted online September 2024. arXiv:2409.16425

22. Van Noorden R. More than 10,000 research papers were retracted in 2023—a new record. Nature. 2023;624(7992):479–481. doi:10.1038/d41586-023-03974-8

23. Fang FC, Steen RG, Casadevall A. Misconduct accounts for the majority of retracted scientific publications. Proc Natl Acad Sci U S A. 2012;109(42):17028–17033. doi:10.1073/pnas.1212247109

24. Freijedo-Farinas F, Ruano-Ravina A, Pérez-Ríos M, Ross J, Candal-Pedreira C. Biomedical retractions due to misconduct in Europe: characterization and trends in the last 20 years. Scientometrics. 2024;129(5):2867–2882. doi:10.1007/s11192-024-04992-7

25. Sridharan K, Sivaramakrishnan G. Artificial intelligence in the retraction spotlight: trends, causes and consequences of withdrawn AI literature through a systematic bibliometric review. Front Res Metr Anal. 2026;10:1737168. doi:10.3389/frma.2025.1737168

26. O’Brien BC, Harris IB, Beckman TJ, Reed DA, Cook DA. Standards for reporting qualitative research: a synthesis of recommendations. Acad Med. 2014;89(9):1245–1251. doi:10.1097/ACM.0000000000000388

27. Reason J. Human Error. Cambridge University Press; 1990.

28. Reason J. Human error: models and management. BMJ. 2000;320(7237):768–770. doi:10.1136/bmj.320.7237.768

29. Landis JR, Koch GG. The measurement of observer agreement for categorical data. Biometrics. 1977;33(1):159–174. doi:10.2307/2529310

30. Macleod MR, Michie S, Roberts I, et al. Biomedical research: increasing value, reducing waste. Lancet. 2014;383(9912):101–104. doi:10.1016/S0140-6736(13)62329-6

31. Moher D, Glasziou P, Chalmers I, et al. Increasing value and reducing waste in biomedical research: who’s listening? Lancet. 2016;387(10027):1573–1586. doi:10.1016/S0140-6736(15)00307-4

32. Carey N, Harte M, Mc Cullagh L. A text-mining tool generated title-abstract screening workload savings: performance evaluation versus single-human screening. J Clin Epidemiol. 2022;149:53–59. doi:10.1016/j.jclinepi.2022.05.017

33. Olofsson H, Brolund A, Hellberg C, et al. Can abstract screening workload be reduced using text mining? User experiences of the tool Rayyan. Res Synth Methods. 2017;8(3):275–280. doi:10.1002/jrsm.1237

34. Macrae C. Managing risk and resilience in autonomous and intelligent systems: exploring safety in the development, deployment, and use of artificial intelligence in healthcare. Risk Anal. 2025;45(4):910–927. doi:10.1111/risa.14273

35. Suchikova Y, Tsybuliak N, Teixeira da Silva JA, Nazarovets S. GAIDeT (Generative AI Delegation Taxonomy): a taxonomy for humans to delegate tasks to generative artificial intelligence in scientific research and publishing. Account Res. 2025;33(3). doi:10.1080/08989621.2025.2544331

