## Supplementary Appendix S1: AI-Assisted Editing Quality-Control Log and Redaction Protocol for "From Output Errors to Workflow Harm: A Practitioner-Audit Method for LLM-Mediated Research"

**AI-Assisted Editing Quality-Control Log and Redaction Protocol**

*Companion to: From Output Errors to Workflow Harm: A Practitioner-Audit Method for LLM-Mediated Research*

Davis Austria, DNP, MSN, MBA, RN, NI-BC, PMP, CPHQ; Byrron McCollister, MS; Jeremy E. Lindsey, MS; Micheal Arowolo, PhD, OCE, MIEEE; Marian Okon, PhD, MPH

MS Health Informatics Program, Xavier University of Louisiana, New Orleans, LA, USA

*Note: This supplement was prepared by the practitioner-author (D.A.) as part of the manuscript's reflexive methodology; co-authors approved its inclusion in the submission package.*

### **1. Purpose**

This supplement provides reviewers with two artifacts. First, it documents specific mistakes the audited system, a frontier commercial conversational LLM, made during the production of the parent manuscript while being used as an editing tool under the author's direction. These mistakes constitute the AI-assisted editing quality-control log referenced in Section 3.3.1 of the parent manuscript. Second, it specifies the redaction protocol the author followed in selecting and de-identifying any quoted material from development conversations, so that the protocol itself is inspectable even where extended verbatim excerpts are not reproduced. The log is provided for reflexive transparency and quality assurance; it is not part of the parent manuscript's primary empirical corpus. This supplement is distinct from Supplementary Appendix S2, which reports the de-identified primary audit corpus (45 incidents) supporting the empirical demonstration.

The supplement does not include a full transcript of the development conversations. The author has chosen to report the audit-relevant mistakes in structured form rather than to reproduce conversational excerpts in this version.

**What this supplement is.** A structured record of mistakes the audited system made during the manuscript's production, with category, description, detection pathway, and correction status for each. The supplement also includes the redaction protocol, the Terms of Service framing for use of conversation materials, and a compliance checklist.

**What this supplement is not.** It is not a behavioral benchmark, a representative sample of audited-system behavior, or a complete record of every editing interaction. It is the author's documentation of audit-relevant mistakes identified through review and external reviewer feedback.

### **2. Redaction protocol**

Where the supplement quotes or paraphrases material from development conversations, redactions follow a three-tier protocol. The protocol is documented here in full so reviewers can evaluate both the standard the author applied and any specific redactions that appear in the documented mistakes table.

#### **2.1 Must be removed**

- Names of collaborators, co-authors, students, reviewers, research assistants, supervisors, or any third party other than the author.
- Specific manuscript identifiers, journal manuscript numbers, grant numbers, IRB application numbers, contract numbers, or project codes.
- Specific dollar figures tied to identifiable incidents beyond what is reported in the parent manuscript at the de-identification level used there.
- Clinical contexts, patient-level information, HIPAA-relevant details, or any content that could identify a clinical encounter.
- Unpublished work titles, working titles, or project identifiers that could identify ongoing research not yet in the public record.
- Personal contact information beyond what is in the manuscript's author block.
- Names of vendors, contractors, business partners, property addresses, legal proceedings, or any third party from contexts outside the audit's scholarly scope.
- Direct content of email, Slack, meeting transcripts, or other private communication referenced in conversation.
- Login credentials, API keys, file paths containing identifying directory names, or system configurations.

#### **2.2 Should be removed**

- Stylistic markers that identify specific collaborators by inference even after their names are removed.
- References to specific institutional processes that could identify the institution beyond what is disclosed in the affiliation block.
- Software versions, file paths, or system configurations beyond what is methodologically necessary.
- Tangential content that is not load-bearing for the methodological point the citation illustrates.

#### **2.3 May stay**

- The author's own positionality references, where the author chooses to surface them. This is the author's editorial decision and is not redacted by default.
- General methodological discussion, citations, and references already present in the parent manuscript.
- References to common tools (Covidence, R, Python, Word, Excel) when no institutional context is attached.
- The author's affiliations and contact information as disclosed in the parent manuscript.

#### **2.4 Redaction marking**

Use square-bracket markers when redacting: [name redacted], [institution redacted], [project redacted], [dollar figure redacted], [details redacted]. Use [...] to indicate trimmed length without substantive removal. No silent deletions.

Before submission the author verified against this protocol that all excerpts in this supplement are de-identified: collaborator and third-party names, institutional and grant identifiers, project codes, and private communication content have been removed, and every redaction is marked with a square-bracket marker rather than silently deleted.

### **3. Terms of Service and consent**

The author treats the conversation materials referenced in this supplement as user-held research materials generated through ordinary use of the audited system. The supplement does not include proprietary system metadata, model weights, internal training documentation, or any non-user-facing information about the audited system's vendor. Because platform terms may change, the author has verified applicable terms at the time of preparation and will comply with journal and institutional requirements regarding use of AI-mediated research materials. Where journal or institutional policy requires additional disclosure or notification beyond what is provided here, the author will provide it before publication.

The audited system's self-descriptions reported in the documented mistakes table (e.g., the system's responses about its own learning or training) are reported as displayed in the conversation. They have not been independently verified against the vendor's published documentation and should be read as the audited system's self-reports rather than as verified architectural facts. This treatment is consistent with the parent manuscript's broader handling of model self-reports.

### **4. Documented editing-interaction mistakes during manuscript production**

Consistent with the parent manuscript's commitment to reflexive transparency about AI-assisted editing, this section documents specific mistakes the audited system made during the production of the manuscript itself, identified through author review of the development conversations and through external reviewer feedback. The author directed the manuscript's argument, structure, interpretation, and final wording; the audited system assisted with editing, synthesis, and prose refinement subject to author verification. The mistakes table below is not exhaustive; it captures the mistakes the author has identified and verified to date. Additional mistakes may emerge during peer review or post-publication, and the author commits to documenting them through the same protocol. The log is provided for reflexive transparency and quality assurance; it is not part of the parent manuscript's primary empirical corpus and is not used to estimate error frequency.

Each mistake is mapped to a category drawn from the parent manuscript's taxonomy. Detection pathway distinguishes errors caught by the author during editing or review, errors caught by external reviewer feedback or reviewer-prompted verification, and errors caught spontaneously by the audited system. Correction status indicates whether the manuscript has been revised, whether a fix is pending, or whether the issue requires further action.

| **ID** | **Category** | **Description** | **Detection pathway** | **Correction status** |
| --- | --- | --- | --- | --- |
| M1 | Claimed verification during editing (citation fabrication) | In manuscript versions v1 through v3, the lead author of the CREOLA paper was cited as "Sapra, A." The actual lead author is Asgari, E. The audited system invented the surname based on no retrieval; the paper had appeared as a URL in earlier search results without an author list being read. | Reviewer-prompted verification. | Corrected in v4 references and Section 2.3 inline. |
| M2 | Claimed verification during editing (citation fabrication) | In manuscript versions v1 through v3, the QualAnalyzer paper was cited as "Ellegood, R., and Blumert, S. (2025)." The actual citation is Lu, M. H., Ellegood, R., Rodriguez-Ramirez, R., & Blumert, S. (2026). The audited system demoted the actual first author (Lu), dropped a co-author (Rodriguez-Ramirez), and assigned the wrong year. Same structural pattern as M1. | Reviewer-prompted verification. | Corrected in v4 references and Section 2.4 inline. |
| M3 | Claimed verification during editing (title fabrication) | HalluLens was cited as "A benchmark for evaluating hallucinations in large language models." The actual title is "HalluLens: LLM Hallucination Benchmark." The audited system produced a descriptive paraphrase that read as the actual title. | Reviewer-prompted verification. | Corrected in v4 references. |
| M4 | Claimed verification during editing (initial fabrication) | The Schwenke et al. autoethnography (Trends in Higher Education, 2023) was cited in manuscript versions v1 through v6 as "Hahn, U. (2023)," with both surname and initial fabricated. The actual lead author is Schwenke, N. The fabrication was not detected until final pre-submission verification. | Reviewer-prompted verification at final pre-submission audit. | Corrected in v7: lead author Schwenke verified, full citation entered with DOI; inline reference in Section 2.2 updated. |
| M5 | Claimed verification during editing (affiliation misattribution) | The audited system characterized the Ibrahim et al. "Beyond Static AI Evaluations" paper as work an industry-aware reviewer would recognize, implying an affiliation that did not exist. Actual affiliations: Oxford Internet Institute, Collective Intelligence Project, OpenAI. | Reviewer-prompted verification. | Did not enter manuscript text; documented here as evidentiary. |
| M6 | Inappropriate framing requiring correction | In a conversational response about the magnitude of the audited system's errors, the audited system invoked the author's positionality (identity-relevant framing) in a discussion where it had no analytic relevance. The framing implicitly suggested that audit credibility depends on the author's identity rather than on the evidence. | Author-flagged in conversation. | Acknowledged. Concrete commitment made not to invoke positionality unless author raises it as analytically relevant. No manuscript text affected. |
| M7 | Regulatory-language overreach | In Section 3.10 of manuscript versions v1 through v3, the language stated that the study "qualifies for IRB exemption under autoethnographic and methodological case-study conventions." Authors do not grant themselves IRB exemption; institutions do. | External reviewer feedback. | Corrected in v5. Section 3.10 now states the author will obtain institutional IRB determination before submission as required by institutional policy. |
| M8 | Regulatory-language overreach | Supplement Section 3 originally contained confident legal posturing about platform terms without institutional or counsel review. | External reviewer feedback. | Corrected in supplement v3. Section 3 language replaced with conservative framing referring to user-held research materials, ordinary use, and institutional/journal policy compliance. |
| M9 | Severity-attribution drift / calibration failure | In manuscript v1, the framing of the $2,500 cost case did partial work toward causal attribution to the audited system without explicit counterfactual analysis. The original language was reasonably conservative but the framing was tighter than the evidence warranted. | Author-flagged during review. | Corrected in v2 with explicit counterfactual paragraph and "necessary but not sufficient" attribution language. |
| M10 | Calibration failure on submission readiness | During v2 and v3 production, the audited system framed remaining work as small editorial improvements when substantial methodological work remained (density check, supplement population, governance language, citation verification). | External reviewer feedback. | Acknowledged. Submission-readiness framing now reflects external reviewer's verdict. |

#### **4.1 Patterns across the documented mistakes**

Five of the ten documented mistakes (M1, M2, M3, M4, M5) are instances of claimed verification during editing applied to citations. This is a clean illustration of the parent manuscript's claimed-verification gap operating in the production of the manuscript itself. The audited system produced citation-formatted output that read as retrieved when retrieval had not occurred or had occurred incompletely. Two (M7, M8) are instances of regulatory-language overreach. One (M6) is inappropriate framing requiring correction. Two (M9, M10) are calibration failures.

The detection-pathway distribution is informative. Eight mistakes were caught only after external reviewer feedback or reviewer-prompted verification (M1, M2, M3, M4, M5, M7, M8, M10). Two were caught by the author during editing or review (M6, M9). Zero were caught by the audited system spontaneously. This is consistent with the parent manuscript's broader argument that practitioner audit, with external review, is the most reliable detection pathway for the failure modes named here. The audited system's correction behavior, when prompted, has been generally specific (naming the failure mode and proposing a concrete remedy) rather than generic apology, but specificity in correction is not equivalent to spontaneous detection.

#### **4.2 Implication for the manuscript's central claim**

The mistakes table provides reflexive quality-control documentation consistent with the manuscript's recommendation for independent verification during AI-assisted scholarly writing. It is not part of the primary empirical corpus and is not used to estimate error frequency. The mistakes table should be interpreted alongside Section 5.5 of the parent manuscript, which frames the editing log as reflexive transparency rather than as part of the primary empirical corpus.

### **5. Notes on use**

This supplement is part of the manuscript submission package. It is referenced in Sections 3.2, 4.7, and 5.2 of the parent manuscript. Reviewers should be able to evaluate the AI participation in manuscript preparation by reading the supplement alongside the parent manuscript, with the documented mistakes table serving as the AI-assisted editing quality-control log.
