## Supplementary Appendix S2: De-identified Audit Corpus Summary for "From Output Errors to Workflow Harm: A Practitioner-Audit Method for LLM-Mediated Research"

**De-identified Audit Corpus Summary**

*Companion to: From Output Errors to Workflow Harm: A Practitioner-Audit Method for LLM-Mediated Research*

Davis Austria, DNP, MSN, MBA, RN, NI-BC, PMP, CPHQ; Byrron McCollister, MS; Jeremy E. Lindsey, MS; Micheal Arowolo, PhD, OCE, MIEEE; Marian Okon, PhD, MPH

MS Health Informatics Program, Xavier University of Louisiana, New Orleans, LA, USA

**Audit corpus:** 45 documented incidents of AI-output error identified during research workflow sessions.

**Documentation window:** March 15, 2026 – May 7, 2026.

**Severity distribution:** 9 critical, 12 high, 18 medium, 6 low.

**Estimated corrective resource impact:** approximately $2,500 (additional reviewer hours, screening restart, third-reviewer adjudication of discrepant records).

**Methodological consequence:** Mid-screening settings changes irreversibly locked records under single-reviewer mode, collapsing the planned dual-review overlap; the resulting κ = 0.224 against the pre-specified κ ≥ 0.70 forced a screening restart on a separate platform.

### **Scope and limitations of this appendix**

This appendix presents a de-identified summary of a longer internal audit log maintained by the practitioner-author across the documentation window. Internal-only details (institution, project names, individual co-author identifiers, and grant numbers) have been removed. The summary preserves date, workflow context, error type, severity, and consequence for each documented incident.

The audit log records confirmed incidents in which AI-generated output required correction, verification, or downstream remediation. It is a record of caught errors and therefore systematically underestimates the true rate of AI output errors during the documentation window. Errors that escaped detection are not, by definition, included.

Final research decisions and analytic interpretations were made by the practitioner-author. Severity ratings reflect the practitioner-author's contemporaneous application of the consequence-based rubric across the documentation window.

This appendix is distinct from Supplementary Appendix S1, which documents AI-assisted editing quality-control issues during preparation of the present manuscript and is not part of the primary empirical corpus reported in this S2.

### **Category normalization**

Raw operational categories assigned at the time of incident documentation were normalized into a smaller set of analytic categories for manuscript-level reporting. Both the raw and normalized categories appear in the table below to permit cross-reference. The manuscript reports incident counts under an eight-category internal taxonomy used in Section 4.1 of the parent manuscript; this appendix uses a finer-grained 13-category normalization. Counts under the two taxonomies do not map one-to-one, and the appendix counts should be read as a more granular view of the same 45 incidents.

**Mapping (raw → normalized):**

| **Raw operational category** | **Normalized analytic category** |
| --- | --- |
| Workflow / Contradictions; Workflow / Safety | Workflow guidance error |
| Factual / Numerical; Factual; Date / Calendar; Meta-Error / Date | Factual error |
| Tool Recommendation; Tool / Platform; Tool / Format; Tool Behavior | Tool / platform misinformation |
| Identity / Positionality | Identity attribution error |
| Unverified Claims; Recommendation Without Verification | Unverified claim presented as fact |
| Behavioral Pattern; Transparency Failure; Severity Communication | Behavioral / interactional pattern |
| Terminology / Construct Error; Citation / Regulatory; Factual / Compliance | Terminology or citation error |
| Citation / XML; Reference Formatting; XML / Tracked Changes; Co-author Edit Acceptance; Co-author List; Text Corruption; Find/Replace Miss | Manuscript-editing error |
| ORCID Misattribution; ORCID Misattribution (chained) | Identifier error |
| Word Count Script | Computation error |
| OCR / False Positive | False-positive error attribution |
| Verification Failure | Verification failure |
| Fabricated Data | Fabrication |
| Financial Consequence | (absorbed into Workflow guidance error) |

### **Severity distribution**

| **Critical** | **High** | **Medium** | **Low** |
| --- | --- | --- | --- |
| 9 | 12 | 18 | 6 |

### **Severity criteria applied**

| **Severity** | **Criterion** |
| --- | --- |
| **Critical** | Methodological breach, fabricated quantitative content in a regulated document, federal-compliance citation error, or incident with documented financial consequence. |
| **High** | Substantive factual error in a manuscript or compliance artifact; identifier or attribution error; unverified claim affecting methodological decision-making. |
| **Medium** | Tool or workflow misinformation requiring rebuild or rework; manuscript-editing error caught pre-submission; behavioral pattern relevant to the analytic frame. |
| **Low** | Cosmetic or low-stakes error caught at a routine review pass; date or formatting inconsistency. |

### **Audit corpus (n = 45)**

*Severity color coding: red = Critical; amber = High; blue = Medium; gray = Low. 'MS example?' indicates whether the incident is referenced as an example in the manuscript text. The audit corpus is presented in two tables for readability: Table A (identification, classification, severity) and Table B (error description, correction pathway, consequence).*

**Summary: incident counts by normalized category**

| **Normalized category** | **n** |
| --- | --- |
| Manuscript-editing error | 10 |
| Tool / platform misinformation | 7 |
| Factual error | 6 |
| Workflow guidance error | 5 |
| Unverified claim presented as fact | 4 |
| Behavioral / interactional pattern | 3 |
| Terminology or citation error | 3 |
| Identifier error | 2 |
| Identity attribution error | 1 |
| Computation error | 1 |
| Verification failure | 1 |
| False-positive error attribution | 1 |
| Fabrication | 1 |
| **Total** | **45** |

*Note on category naming. The "Manuscript-editing error" category refers to manuscript-development incidents captured in the original 45-incident audit corpus during the March 15 to May 7, 2026 documentation window in the practitioner-author’s prior scholarly work. It does not include the separate AI-assisted editing quality-control observations generated during preparation of the present manuscript, which are documented in Supplement S1 and are not part of the primary empirical corpus.*

*Note: This 13-category normalization is finer-grained than the 8-category internal taxonomy used in Section 4.1 of the parent manuscript. Counts under the two taxonomies are parallel views of the same 45 incidents and do not map one-to-one.*

**Table A. Incident identification, classification, and severity**

| **ID** | **Date** | **Workflow** | **Raw category** | **Normalized category** | **Severity** |
| --- | --- | --- | --- | --- | --- |
| 1 | 2026-03-17/24 | Scoping review screening configuration | Workflow / Contradictions | Workflow guidance error | **Critical** |
| 2 | 2026-03-17/24 | Scoping review IRR computation | Factual / Numerical | Factual error | **High** |
| 3 | 2026-03-17/24 | Scoping review tool selection | Tool Recommendation | Tool / platform misinformation | **High** |
| 4 | 2026-03-17/24 | Database export workflow | Tool Recommendation | Tool / platform misinformation | **Medium** |
| 5 | 2026-03-17/24 | Search strategy construction | Tool Recommendation | Tool / platform misinformation | **Medium** |
| 6 | 2026-03-17/24 | Manuscript drafting (positionality) | Identity / Positionality | Identity attribution error | **High** |
| 7 | 2026-03-17/24 | Screening interface guidance | Tool / Platform | Tool / platform misinformation | **Low** |
| 8 | 2026-03-17/24 | Screening platform behavior | Tool Behavior | Tool / platform misinformation | **Medium** |
| 9 | 2026-03-17/24 | Reference manager workflow | Tool Behavior | Tool / platform misinformation | **Low** |
| 10 | 2026-03-17/24 | Sprint planning | Date / Calendar | Factual error | **Low** |
| 11 | 2026-04-26 | Audit documentation (meta) | Meta-Error / Date | Factual error | **Low** |
| 12 | 2026-04-26 | Screening platform settings change | Workflow / Safety | Workflow guidance error | **Critical** |
| 13 | 2026-04-26 | Data integrity verification | Unverified Claims | Unverified claim presented as fact | **High** |
| 14 | 2026-04-26 | Platform feature description | Unverified Claims | Unverified claim presented as fact | **Medium** |
| 15 | 2026-04-26 | Cross-platform data transfer | Tool / Format | Tool / platform misinformation | **Medium** |
| 16 | 2026-04-26 | Cross-cutting interaction pattern | Behavioral Pattern | Behavioral / interactional pattern | **Medium** |
| 17 | 2026-04-26 | AI self-explanation | Transparency Failure | Behavioral / interactional pattern | **Medium** |
| 18 | 2026-04-26 | Manuscript methodology | Recommendation Without Verification | Unverified claim presented as fact | **Medium** |
| 19 | 2026-04-27 | Construct definition (Project A) | Terminology / Construct Error | Terminology or citation error | **High** |
| 20 | 2026-04-27 | Conference submission guidance | Factual | Factual error | **Medium** |
| 21 | 2026-04-27 | Credentialing compliance | Factual / Compliance | Terminology or citation error | **Medium** |
| 22 | 2026-05-04/05 | Scoping review IRR computation | Factual / Numerical | Factual error | **High** |
| 23 | 2026-05-04/05 | Data integrity verification | Unverified Claims | Unverified claim presented as fact | **High** |
| 24 | 2026-05-04/05 | Screening platform settings change | Workflow / Safety | Workflow guidance error | **Critical** |
| 25 | 2026-05-04/05 | Scoping review screening configuration | Workflow / Contradictions | Workflow guidance error | **High** |
| 26 | 2026-05-04/05 | Cross-cutting consequence | Financial Consequence | Workflow guidance error | **Critical** |
| 27 | 2026-05-04/05 | Severity communication | Severity Communication | Behavioral / interactional pattern | **Critical** |
| 28 | 2026-04/05 | Manuscript word-count audit | Word Count Script | Computation error | **Medium** |
| 29 | 2026-04/05 | Manuscript citation placement | Citation / XML | Manuscript-editing error | **Medium** |
| 30 | 2026-04/05 | Manuscript reference formatting | Reference Formatting | Manuscript-editing error | **Medium** |
| 31 | 2026-04/05 | Manuscript reference numbering | Citation / XML | Manuscript-editing error | **Medium** |
| 32 | 2026-04/05 | Manuscript tracked-changes acceptance | XML / Tracked Changes | Manuscript-editing error | **Medium** |
| 33 | 2026-04/05 | Manuscript tracked-changes acceptance | Co-author Edit Acceptance | Manuscript-editing error | **High** |
| 34 | 2026-04/05 | Manuscript co-author list | Co-author List | Manuscript-editing error | **High** |
| 35 | 2026-04/05 | Manuscript text insertion | Text Corruption | Manuscript-editing error | **Critical** |
| 36 | 2026-04/05 | Identifier management | ORCID Misattribution | Identifier error | **High** |
| 37 | 2026-04/05 | File verification | Verification Failure | Verification failure | **Critical** |
| 38 | 2026-04/05 | Manuscript find/replace | Find/Replace Miss | Manuscript-editing error | **Medium** |
| 39 | 2026-04/05 | Manuscript find/replace | Find/Replace Miss | Manuscript-editing error | **Medium** |
| 40 | 2026-04/05 | Identifier management | ORCID Misattribution (chained) | Identifier error | **High** |
| 41 | 2026-04/05 | Manuscript reference formatting | Reference Formatting | Manuscript-editing error | **Low** |
| 42 | 2026-04/05 | Figure review (OCR-mediated) | OCR / False Positive | False-positive error attribution | **Medium** |
| 43 | 2026-04/05 | Submission guidance | Factual | Factual error | **Low** |
| 44 | 2026-05-07 | Accreditation self-study drafting | Fabricated Data | Fabrication | **Critical** |
| 45 | 2026-05-07 | IRB application drafting | Citation / Regulatory | Terminology or citation error | **Critical** |

**Table B. Incident description, correction pathway, consequence, and manuscript-example flag**

| **ID** | **Error summary** | **Correction source** | **Consequence** | **MS example?** |
| --- | --- | --- | --- | --- |
| 1 | AI output provided multiple internally contradictory instructions on screening-platform reviewer settings within a single session. | Practitioner-author-initiated re-verification with vendor support | Mid-screening settings change locked records under single-reviewer mode, collapsing planned reviewer overlap (resulting κ = 0.224 vs pre-specified κ ≥ 0.70); estimated corrective resource impact ~$2,500. | Yes (primary case) |
| 2 | AI output reported inconsistent figures for the IRR overlap sample across the same session. | Direct vendor export by practitioner-author | Required practitioner-author to obtain authoritative figures from platform support to resolve discrepancy. | Yes |
| 3 | AI recommended a screening platform without first assessing capacity limits, deduplication options, or institutional access. | Practitioner-author discovery during attempted use | Time loss to failed uploads and workarounds before transitioning to the institutionally supported platform. | No |
| 4 | AI recommended a database export workflow without anticipating known per-batch record limits. | Practitioner-author discovery during attempted export | Workflow rebuild required; correct conversion path identified after delay. | No |
| 5 | AI built a database into a five-source search strategy without verifying institutional access tier. | Practitioner-author-initiated institutional verification | Search strategy rebuilt; alternative database substituted. | No |
| 6 | AI drafted a positionality statement misattributing the practitioner-author's racial/ethnic identity. | Practitioner-author correction at draft review | Statement required correction; affects equity framing of the work. | Yes |
| 7 | AI provided keyboard shortcuts that did not function on the practitioner-author's operating system in the screening platform. | Practitioner-author discovery during screening | Screening guide required rewrite for the correct interaction model. | No |
| 8 | AI implied automated filtering behavior in a screening platform that in fact requires manual decisions. | Practitioner-author-initiated clarification | False expectation corrected only after direct query. | No |
| 9 | AI recommended a reference-manager merger workflow without warning of dual-collection import behavior. | Practitioner-author discovery via inflated counts | Troubleshooting required to reconcile record counts. | No |
| 10 | AI output contained day-of-week / date mismatches in sprint planning artifacts. | Practitioner-author correction at calendar check | Sprint plan dates corrected; flagged as recurring pattern. | No |
| 11 | Within an internal audit document, AI output stated an incorrect day-of-week as part of a correction to a prior date error. | Practitioner-author second-pass audit | Correction-of-correction required. | No |
| 12 | AI advised a configuration change without proposing pre-change safeguards (export, screenshot, spot-check, rollback). | Practitioner-author comparison with alternate AI guidance | No backup existed when the platform displayed an irreversibility warning during the change. | Yes |
| 13 | AI asserted that data integrity could be confirmed from screenshots when the platform did not support such verification. | Practitioner-author-initiated direct platform export | False confidence influenced subsequent decisions. | Yes |
| 14 | AI stated a specific record-count threshold for a platform feature without documentation review. | Practitioner-author-initiated documentation check | Unverified technical claim presented as fact during methodological decision-making. | No |
| 15 | AI stated that a screening platform accepts a file format it does not support. | Practitioner-author discovery in target platform | Practitioner-author independently identified correct conversion path. | No |
| 16 | AI output across multiple sessions used language directing the practitioner-author to end the conversation or postpone follow-up. | Practitioner-author cross-session pattern analysis | Identified as a candidate trained-behavior pattern relevant to the manuscript's analytic frame. | Yes |
| 17 | AI output expressed greater certainty than warranted when asked to characterize its own behavioral patterns. | Practitioner-author direct query | Inability to provide an authoritative account of underlying mechanisms when queried; relevant to the manuscript's transparency analysis. | Yes |
| 18 | AI presented and recommended a screening approach without first verifying target-journal author guidelines. | Practitioner-author-initiated guideline check | Recommendation should not have been made absent verification. | No |
| 19 | AI misnamed a foundational measurement construct and generated nonexistent subdomains. | Practitioner-author construct verification | Core construct error caught pre-submission; would have undermined framework credibility in peer review. | Yes |
| 20 | AI stated a professional society did not have a relevant submission category when one existed. | Practitioner-author direct verification with source | Incorrect submission guidance; caught before action. | No |
| 21 | AI mischaracterized the scope of a professional credentialing disclosure section. | Practitioner-author direct verification with source | Incorrect compliance guidance; would have led to unnecessary disclosures on a credentialing document. | No |
| 22 | AI accepted or computed an IRR overlap value approximately one-ninth of the actual export figure. | Practitioner-author-initiated platform export | Substantively understated the IRR sample, affecting the accuracy of subsequent guidance. | Yes |
| 23 | AI asserted that the IRR sample was intact and clean based on dashboard screenshot interpretation. | Practitioner-author-initiated platform export | Severity of methodological breach was not communicated until practitioner-author obtained authoritative export. | Yes |
| 24 | AI advised a settings change without flagging the platform's explicit irreversibility warning. | Platform-displayed warning encountered by practitioner-author | Contributed to records being locked under single-reviewer mode. | Yes |
| 25 | AI output continued a within-session pattern of contradictory configuration guidance. | Practitioner-author-initiated vendor support consultation | Practitioner-author obtained authoritative guidance from platform support. | Yes |
| 26 | Cumulative AI-output errors across sessions were associated with the need for a screening restart, additional reviewer hours, and adjudication of discrepant records. | Practitioner-author accounting of corrective actions | Estimated corrective resource impact ~$2,500; schedule impact including delayed funder deliverables, website development, and an IRB submission. | Yes (primary case) |
| 27 | AI output minimized the severity of the IRR result and did not flag immediate co-author notification as the appropriate next step. | Practitioner-author judgement | Co-author notification was delayed. | Yes |
| 28 | AI word-count script included end matter in body word count. | Practitioner-author-initiated re-computation | Unnecessary trimming discussion; corrected pre-submission. | No |
| 29 | AI attached a citation superscript to an adjacent self-referential sentence rather than the supporting findings sentence. | Practitioner-author manual correction | Citation displacement requiring manual repair. | No |
| 30 | AI programmatic builds did not apply required italic formatting for journal names per AMA 11th Edition. | Practitioner-author manual correction in Word | Reference list required full manual correction. | No |
| 31 | AI renumbering script double-incremented a hardcoded reference number, displacing subsequent reference assignments. | Practitioner-author reference-list audit | Targeted XML repair required. | No |
| 32 | AI acceptance of co-author tracked changes left split XML runs in a citation cluster. | Practitioner-author XML inspection | Citation rendering corruption requiring XML-level repair. | No |
| 33 | AI accepted a substantive co-author edit ('proposed study' for an active study) without flagging it as a decision point. | Practitioner-author manual review | Substantive content error caught and reverted pre-submission. | No |
| 34 | AI accepted a co-author version of the manuscript missing one named co-author without flagging the absence. | Practitioner-author verification audit | Near-miss; missing co-author re-inserted before submission. | No |
| 35 | AI sentence insertion produced a corrupted residual fragment in a Methods paragraph. | External reviewer detection in subsequent review round | Most consequential manuscript-editing incident; escaped internal audit until detected externally. | Yes |
| 36 | AI output used an incorrect researcher identifier across multiple sessions and downstream artifacts. | Practitioner-author correction | Identity error on a manuscript-tracking artifact. | No |
| 37 | AI applied fixes to one file location and verified them against a different cached copy rather than the delivered output. | Practitioner-author push-back for re-verification | Delivered output retained unfixed content until practitioner-author-initiated second check. | Yes |
| 38 | AI find/replace pass missed an instance of a deprecated term in an appendix. | Practitioner-author-initiated cross-file second check | Inconsistent terminology across submission package; corrected pre-submission. | No |
| 39 | AI find/replace pass missed multiple instances of deprecated terms in manuscript end matter; verification was performed against a cached file. | Practitioner-author-initiated cross-file second check | Inconsistent terminology corrected pre-submission. | No |
| 40 | During correction of a prior identifier error, AI output reattributed the incorrect identifier to a second co-author, also incorrectly. | Practitioner-author cross-check against authoritative records | Chained error during a correction action; resolved by practitioner-author. | No |
| 41 | AI omitted the 'Accessed' qualifier preceding an access date in one reference, inconsistent with adjacent references. | Practitioner-author final spot check | AMA-style citation inconsistency caught pre-submission. | No |
| 42 | AI OCR-based review of figure images produced misreadings that were reported as figure errors when the underlying figures were correct. | Practitioner-author push-back and direct visual inspection | Wasted review cycles; figures confirmed correct. | No |
| 43 | AI stated a target-journal article processing fee that exceeded the actual fee on the submission completion page. | Practitioner-author verification on submission page | Inaccurate budget guidance. | No |
| 44 | AI output included a specific quantitative enrollment figure in self-study draft content that was not supported by source institutional records. | Practitioner-author source-document verification | High-stakes accreditation document near-miss; unsupported quantitative claim removed pre-submission. | Yes |
| 45 | AI output cited a federal regulatory subpart in an IRB application context where the subpart was misapplied to the protocol's scope. | Practitioner-author regulatory verification | Caught pre-submission; would have introduced an incorrect regulatory citation into a federal-compliance document. | Yes |

### **Note on this appendix**

This summary is derived from a longer internal audit log retained by the practitioner-author. The internal log contains additional operational detail (specific tool screenshots, exact wording of AI output, internal communication notes) that is not appropriate for the public record. The de-identified summary above preserves the analytic content needed to evaluate the manuscript's claims regarding the audit corpus.

**Addendum: 13-to-8-category crosswalk and author coding record (added July 2026)**

The parent manuscript reports incident counts under a locked eight-category taxonomy; this appendix uses the finer-grained 13-category normalization above. The empirical mapping between the two, derived from the practitioner-author's per-incident coding of all 45 incidents, is shown below. Seven of the 13 normalized categories split across manuscript categories; the mapping is many-to-many by design and is governed by the coding conventions listed after the table.

**Operational definitions of the eight manuscript categories:**

| **Manuscript category** | **Operational definition** |
| --- | --- |
| Verification failure | False or implied performance of a verification process: the output claims or implies that a check occurred, or asserts direct access to an artifact's state, when no adequate check took place. |
| Factual numerical error | Wrong number, date, value, or fabricated quantitative content. |
| Tool-behavior misunderstanding | Misrepresentation of platform, tool, format, or system behavior or capability. |
| Unverified claim presented as fact | Unsupported factual assertion without an explicit or implied claim that verification occurred. |
| Citation or reference formatting error | Citation, reference, terminology, or regulatory-citation defect, including reference-apparatus omissions. |
| Workflow contradiction | Contradictory or shifting procedural guidance, including loss of prior task constraints. |
| Document or protocol-safety failure | Structural document-integrity damage or unsafe protocol guidance. |
| Identity or positionality | Misassigned identity, attribution, or identifier. |

| **S2 normalized category (n)** | **Manuscript 8-category assignment(s)** |
| --- | --- |
| Manuscript-editing error (10) | 5× Citation or reference formatting error; 3× Document or protocol-safety failure; 2× Verification failure |
| Tool / platform misinformation (7) | 7× Tool-behavior misunderstanding |
| Factual error (6) | 5× Factual numerical error; 1× Verification failure |
| Workflow guidance error (5) | 3× Workflow contradiction; 2× Document or protocol-safety failure |
| Unverified claim presented as fact (4) | 3× Verification failure; 1× Unverified claim presented as fact |
| Behavioral / interactional pattern (3) | 2× Unverified claim presented as fact; 1× Workflow contradiction |
| Terminology or citation error (3) | 2× Citation or reference formatting error; 1× Document or protocol-safety failure |
| Identifier error (2) | 2× Identity or positionality |
| Identity attribution error (1) | 1× Identity or positionality |
| Computation error (1) | 1× Factual numerical error |
| Verification failure (1) | 1× Verification failure |
| False-positive error attribution (1) | 1× Unverified claim presented as fact |
| Fabrication (1) | 1× Factual numerical error |

Resulting manuscript-taxonomy counts (n=45): verification failure 7; factual numerical error 7; tool-behavior misunderstanding 7; citation or reference formatting error 7; document or protocol-safety failure 6; workflow contradiction 4; unverified claim presented as fact 4; identity or positionality 3.

**Coding conventions (recorded during adjudication):**

• Process test: an incident is Verification failure when the output claims or implies that a verification act occurred, or asserts direct access to an artifact's state, regardless of whether the content is also wrong; a false process claim takes precedence over the content error.

• Unverified claim covers assertions stated as fact with no process claim; a check that occurred but was performed defectively is coded by the resulting assertion, not as Verification failure.

• Silent acceptance of a substantive content change into a deliverable implies that review occurred and is coded Verification failure; mechanical artifacts of an acceptance operation are Document or protocol-safety failure; omissions in reference apparatus (including author lists) are Citation or reference formatting error, while misassigned identities and identifiers are Identity or positionality.

• Structural corruption of a document (truncation, residual fragments, split runs) is Document or protocol-safety failure even when it arises inside citation content.

Per-incident manuscript-taxonomy assignments for the full corpus were finalized by the practitioner-author in July 2026 under the conventions above; the sixteen-incident subsample coding is unchanged from the June 2026 audit trail, and the complete coding record is retained in the project audit trail.
