## Supplementary Appendix S3: Audit-Log Coding Schema and User Agency Protocol for "From Output Errors to Workflow Harm: A Practitioner-Audit Method for LLM-Mediated Research"

**Audit-Log Coding Schema and User Agency Protocol**

*Companion to: From Output Errors to Workflow Harm: A Practitioner-Audit Method for LLM-Mediated Research*

Davis Austria, DNP, MSN, MBA, RN, NI-BC, PMP, CPHQ; Byrron McCollister, MS; Jeremy E. Lindsey, MS; Micheal Arowolo, PhD, OCE, MIEEE; Marian Okon, PhD, MPH

MS Health Informatics Program, Xavier University of Louisiana, New Orleans, LA, USA

**1. Purpose**

This appendix provides two operational instruments of the TRACE practitioner-audit method that are referenced in the main manuscript but held here to keep the article within journal table limits: the audit-log coding schema (the per-incident data structure used to record each incident) and the User Agency Protocol (the practitioner practices that convert transient AI interactions into auditable evidence). The Response-Audit Scorecard is presented in the main text (Table 3).

**2. Audit-log coding schema**

Each documented incident was recorded using the fields below. The schema is the data structure underlying the de-identified corpus summarized in Supplementary Appendix S2.

| **Field** | **Description** | **Example coding options** |
| --- | --- | --- |
| Incident ID | Unique identifier and date | C-YYYYMMDD-## |
| Model label | Model name as displayed or recorded | Audited system, version as displayed; secondary system if used; unknown if not visible |
| Workflow context | Work area affected | Scoping review; manuscript; IRB; credentialing; program review |
| User task | What the model was asked to do | Draft, verify, summarize, calculate, revise, format, troubleshoot |
| Model behavior | What the model did | Fabricated value; contradicted prior instruction; claimed verification; truncated text |
| Error definition | Primary error type | Factual; unsupported; false verification; context contradiction; tool misunderstanding; document integrity; response inconsistency |
| Detection pathway | How the issue was found | Practitioner review; external reviewer; source check; document comparison; downstream failure |
| Correction source | Evidence used to confirm the error | Authoritative source; institutional record; manuscript comparison; protocol; software behavior |
| Downstream consequence | What happened or could have happened | Rework; submission risk; reporting risk; screening disruption; trust recalibration |
| Severity | Consequence-based severity | Critical; high; medium; low |
| Taxonomy mapping | Research category alignment | Factuality hallucination; faithfulness failure; calibration; false refusal; sycophancy; long-context failure; claimed verification |
| Response after correction | How the assistant responded | Direct acknowledgment; generic apology; defensive framing; concrete safeguard; commitment failure |
| Recurrence link | Whether similar errors appeared before or after | Related incident IDs or category recurrence |

**3. User Agency Protocol**

The User Agency Protocol is the set of practitioner practices that make AI-mediated workflow interactions auditable. It spans the workflow-harm pathway stages and is the active response infrastructure of the method.

| **Practice** | **Purpose** |
| --- | --- |
| Freeze the output | Preserve the exact prompt, response, date, model label, and affected file before the interaction changes. |
| Require an incident record | Convert the AI mistake into a structured account with error type, correction source, harm, severity, and safeguard. |
| Apply a stop-work rule | Prevent AI-generated procedural, regulatory, statistical, citation, or software guidance from being used until externally verified. |
| Keep a cost-of-error ledger | Track funds spent, added software, rework hours, delayed milestones, and unrecoverable labor time. |
| Audit the AI response | Score whether the AI's correction behavior reduces recurrence risk or merely apologizes. |
| Link recurrence | Connect similar errors across sessions to show pattern, not isolated inconvenience. |
| De-identify for publication | Remove patient, student, collaborator, reviewer, institutional, and unpublished project identifiers before dissemination. |
| Persist the safeguard | Convert session-level corrections into standing, program-level controls—workspace convention files, standing-correction documents, and reusable enforcement skills that load automatically in subsequent sessions—so prevention does not depend on per-session practitioner recall or re-instruction. |

**4. Provenance**

- **Companion manuscript:** From Output Errors to Workflow Harm: A Practitioner-Audit Method for LLM-Mediated Research.
- **Related appendices:** S1 (AI-assisted editing quality-control log); S2 (de-identified audit corpus summary, n = 45).
- Moved from the main text for JAMIA table-limit compliance; the Response-Audit Scorecard remains in the body as Table 3.
