## Supplementary Appendix S4: Replication Workflow for Practitioner Audits for "From Output Errors to Workflow Harm: A Practitioner-Audit Method for LLM-Mediated Research"

**Replication Workflow for Practitioner Audits**

*Companion to: From Output Errors to Workflow Harm: A Practitioner-Audit Method for LLM-Mediated Research*

Davis Austria, DNP, MSN, MBA, RN, NI-BC, PMP, CPHQ; Byrron McCollister, MS; Jeremy E. Lindsey, MS; Micheal Arowolo, PhD, OCE, MIEEE; Marian Okon, PhD, MPH

MS Health Informatics Program, Xavier University of Louisiana, New Orleans, LA, USA

**1. Purpose**

This appendix provides a step-by-step workflow for researchers who wish to replicate the TRACE practitioner-audit method in their own scholarly, clinical informatics, or public-health workflows. It is designed to be executable using only user-facing conversational interfaces, de-identified logs, and practitioner documentation, without vendor-internal access or proprietary system metadata.

**2. Prerequisites**

- An ongoing workflow in which an LLM assistant is used for consequential scholarly or clinical-adjacent tasks.
- The four TRACE instruments: the severity rubric (manuscript Table 1), the locked 8-category taxonomy (manuscript Section 4.1; operational definitions in Supplementary Appendix S2), the Response-Audit Scorecard (manuscript Table 3), and the audit-log coding schema and User Agency Protocol (Supplementary Appendix S3).
- A redaction/de-identification protocol (Supplementary Appendix S1).

**3. Step-by-step workflow**

**Phase 0 — Set up audit infrastructure**

- Adopt the audit-log coding schema (Supplementary Appendix S3) as the per-incident data structure. Apply the User Agency Protocol, beginning with the freeze-the-output practice: preserve the exact prompt, response, date, model label, and affected file before the interaction changes.

**Phase 1 — Define what counts as an incident**

- Adopt the error definition (manuscript Section 3.4): log an incident when output or interaction meets at least one criterion (factual/numerical, regulatory, or bibliographic inaccuracy; unsupported claim as fact; claimed-but-absent verification; context or workflow contradiction; tool/system misunderstanding; document-integrity failure; or refusal/response-pattern inconsistency).
- Use a consequence-driven documentation threshold; expect a documentation-positive subset, not a full error census.

**Phase 2 — Document incidents**

- For each incident, record the schema fields: identifier and date, workflow context, model behavior, downstream consequence, correction source, and notes.

**Phase 3 — Code severity and category**

- Assign consequence-based severity (Critical/High/Medium/Low; Table 1) and a category from the locked 8-category taxonomy, plus a claimed-verification flag (Yes/No/Unclear).

**Phase 4 — Score post-error response**

- Apply the Response-Audit Scorecard (manuscript Table 3) to the assistant's behavior after correction across the seven criteria.

**Phase 5 — De-identify**

- Apply the three-tier redaction protocol (Supplementary Appendix S1) before any material leaves the private audit trail.

**Phase 6 — Independent reviewer coding check**

- Draw a stratified random subsample (record the seed and selected IDs). Have two or more independent reviewers blind-code category, severity, and the claimed-verification flag. Compute Cohen's kappa (nominal) and weighted kappa (ordinal severity). Report agreement honestly; low agreement is a finding about taxonomization difficulty, not a defect to be re-coded away.

**Phase 7 (optional) — AI-comparator taxonomy-interpretability check**

- Have three or more structurally distinct LLMs independently apply the taxonomy to the full incident set, at temperature 0, with the instrument embedded verbatim in the prompt and each system blinded. Compute Fleiss's kappa and pairwise Cohen's kappa across systems. Frame results as taxonomy legibility, not reliability; exclude any system used elsewhere in the study to avoid circularity.

**Phase 8 — Analyze and report**

- Report per SRQR. Argue adequacy from saturation, not enumeration. Report the workflow-harm patterns, the claimed-verification finding, and the response-audit findings.

**4. Reproducibility parameters (AI-comparator phase)**

- **Comparators:** three independent, structurally distinct frontier LLM comparators from different developer lineages, run at temperature 0 (with deterministic seeding fixed where a system exposes it). Exact model identifiers and run parameters are retained in the study audit trail and are editor-available, consistent with the vendor-blinding used in the manuscript and Supplementary Appendix S5.
- **Instrument delivery:** the full taxonomy and rubrics are embedded verbatim in the system prompt; models are asked to output the category label text only, with no leading number.
- **Analysis normalization:** category labels compared case-insensitively with any leading enumeration prefix stripped.

**5. Cautions**

- The corpus is documentation-positive; counts are not frequency estimates.
- AI-comparator agreement is a legibility signal, not reliability; convergence may partly reflect shared model priors.
- Model labels reflect the practitioner's interface, not verified system metadata; do not read them as vendor comparisons.
