## Supplementary Appendix S5: AI-Comparator Analysis Materials and Results for "From Output Errors to Workflow Harm: A Practitioner-Audit Method for LLM-Mediated Research"

**AI-Comparator Analysis Materials and Results**

*Companion to: From Output Errors to Workflow Harm: A Practitioner-Audit Method for LLM-Mediated Research*

Davis Austria, DNP, MSN, MBA, RN, NI-BC, PMP, CPHQ; Byrron McCollister, MS; Jeremy E. Lindsey, MS; Micheal Arowolo, PhD, OCE, MIEEE; Marian Okon, PhD, MPH

MS Health Informatics Program, Xavier University of Louisiana, New Orleans, LA, USA

**1. Purpose and framing**

This appendix provides the materials and results for the supplemental AI-comparator taxonomy-interpretability analysis referenced in the manuscript (Sections 3.12 and 4.8). Its purpose is reproducibility of the method and of the reported agreement values.

*This analysis assesses taxonomy interpretability (cross-system legibility), not reliability. It is not a validation of the incidents and does not revise or replace the human reviewer-author coding.*

**2. Comparators**

- **Comparators:** three independent, structurally distinct frontier LLM comparators from different developer lineages, accessed via hosted APIs. The audited conversational system (used for manuscript editing) was excluded from the comparator set to avoid circularity and is vendor-blinded in the manuscript.
- **Run parameters:** temperature 0 for all comparators; where a system exposed deterministic seeding it was fixed, otherwise outputs retain run-to-run stochasticity; the instrument was delivered verbatim in the system prompt.
- **Exact identifiers:** exact model identifiers, versions, endpoints, and run parameters are retained in the study audit trail and can be provided to editors for review, reproducibility checking, or post-publication disclosure if required by journal policy.

**3. Instrument (delivered verbatim to each comparator)**

**3.1 Role**

Each comparator was instructed to act as a blinded independent comparator applying a fixed incident-coding taxonomy, to code each incident independently, and not to revise earlier codings after coding later ones. It was told its outputs are used to assess taxonomy legibility and that it is not an author, reviewer, or researcher on the study.

**3.2 Category taxonomy (8 codes; primary analysis)**

Choose exactly one; output the category label text only, with no leading number.

- verification failure
- factual numerical error
- tool-behavior misunderstanding
- unverified claim presented as fact
- citation or reference formatting error
- workflow contradiction
- document or protocol-safety failure
- identity or positionality

**3.3 Severity, claimed-verification, confidence**

- **Severity (one of):** Critical / High / Medium / Low, by workflow consequence.
- **Claimed-verification flag (one of):** Yes / No / Unclear.
- **Confidence:** 0.0 to 1.0, descriptive only (not a calibrated probability or validity weight).

**3.4 Response format**

One JSON object per incident with fields: incident_id, category_code, severity_code, claimed_verification, confidence, methodological_note. No text outside the JSON object.

**3.5 Category normalization (analysis side)**

Category labels were compared case-insensitively, with any leading enumeration prefix (for example, “8. ”) stripped, before computing agreement. Severity and the claimed-verification flag share casing and were not normalized.

**4. Primary results: 8-category, all 45 incidents**

Comparators are labeled A, B, and C (fixed within this appendix; the mapping to exact identifiers is in the audit trail).

| **Agreement** | **Dimension** | **Kappa** | **Landis-Koch** |
| --- | --- | --- | --- |
| Fleiss (A+B+C) | Category | +0.632 | substantial |
| Fleiss (A+B+C) | Severity | +0.401 | fair |
| Fleiss (A+B+C) | Claimed-verification | +0.405 | fair |
| Pairwise Cohen | Category, A–B | +0.615 | substantial |
| Pairwise Cohen | Category, A–C | +0.759 | substantial |
| Pairwise Cohen | Category, B–C | +0.536 | moderate |

**5. Human anchor (16-incident subsample)**

On the 16-incident stratified subsample coded by the three human reviewer-authors, independent category agreement was slight (Fleiss κ = 0.155), with the claimed-verification flag at κ = 0.005; comparator–human differences reflect taxonomy legibility under standardized conditions, not superior comparator judgment. Cross-comparator category agreement (0.632 on the full corpus) exceeded this, the manuscript's central legibility signal. Comparator ratings aligned more closely with one reviewer-author than the other, mirroring the human-to-human asymmetry.

**6. Sensitivity: 10-category instrument, all 45 incidents**

A standalone sensitivity analysis added two reviewer-motivated categories (identifier error; behavioral or interactional pattern). Category-level agreement did not fall and was somewhat higher (Fleiss κ = 0.690); severity (0.364) and claimed-verification (0.363) remained fair. All three comparators assigned the added categories predominantly to the incidents that motivated them. Because the 8- and 10-category runs are separate and unpaired, the result is read as “did not reduce category-level interpretability,” not as a demonstrated improvement.

**7. Interpretation and cautions**

- Cross-comparator agreement is a taxonomy-legibility signal, not a reliability statistic, and not a validation of the codings.
- Because the comparators may share overlapping training data and architecture, their agreement may partly reflect shared model priors; the analysis is triangulation against, not confirmation of, the human coding.
- Full analysis scripts, prompts logs, parsed codings, and kappa reports are retained in the study audit trail.
